# Multivariate Genomic Architecture of Cortical Thickness and Surface Area at Multiple Levels of Analysis

**DOI:** 10.1101/2022.02.19.22271223

**Authors:** Andrew D. Grotzinger, Travis T. Mallard, Zhaowen Liu, Jakob Seidlitz, Tian Ge, Jordan W. Smoller

**Affiliations:** Institute for Behavioral Genetics, University of Colorado Boulder, Boulder, CO USA; Department of Psychology and Neuroscience, University of Colorado Boulder, Boulder, CO USA; Psychiatric and Neurodevelopmental Genetics Unit, Massachusetts General Hospital, Boston, MA; Center for Precision Psychiatry, Department of Psychiatry, Massachusetts General Hospital, Boston, MA; Stanley Center for Psychiatric Research, Broad Institute, Cambridge, MA; Department of Psychiatry, University of Pennsylvania, Philadelphia, PA; Department of Child and Adolescent Psychiatry and Behavioral Science, The Children’s Hospital of Philadelphia, Philadelphia, PA

**Author notes:** Correspondence to Andrew D. Grotzinger. These authors contributed equally to this manuscript (Travis T. Mallard, Zhaowen Liu).

## Abstract

Recent work in imaging genetics suggests high levels of genetic overlap within cortical regions for cortical thickness (CT) and surface area (SA). We model this multivariate system of genetic relationships by applying Genomic Structural Equation Modeling (Genomic SEM) to parsimoniously define five genomic brain factors for both CT and SA. We reify these factors by demonstrating the generalizability of the model in a semi-independent sample and show that the factors align with biologically and functionally relevant parcellations of the cortex. We apply Stratified Genomic SEM to identify specific categories of genes (e.g., neuronal cell types) that are disproportionately associated with pleiotropy across specific subclusters of brain regions, as indexed by the genomic factors. Finally, we examine genetic associations with psychiatric and cognitive correlates, finding that SA is associated with both broad aspects of cognitive function and specific risk pathways for psychiatric disorders. These analyses provide key insights into the multivariate genomic architecture of two critical features of the cerebral cortex.

---

The human cerebral cortex broadly refers to the outer sheet of gray matter and is typically indexed using two key metrics: cortical thickness (CT) and surface area (SA). In practice, CT is operationalized as the distance between pial surfaces and white matter, and SA as geodesics along the grey-white matter boundary. These two measures are both key predictors of important life outcomes, with CT generally associated with a range of psychiatric disorders,^1,2^ and SA with a host of cognitive outcomes across the lifespan.^3–6^ In the last decade, twin studies have shown that these two metrics are highly heritable, while characterized by distinct genetic underpinnings.^7,8^ Even more recently, genotyped samples with neuroimaging data are now large enough to employ genome-wide association studies (GWAS) as a means of identifying the specific genetic variants associated with these structural outcomes. For example, the ENIGMA consortium examined bilateral averages of 34 cortical brain regions to identify 175 and 48 genetic loci associated with regional SA and CT, respectively.^9^ Two additional findings include the observation that, consistent with the family-based literature, the different SA and CT brain regions were highly positively correlated within measures of CT and SA and negatively correlated across CT and SA.^9^ These results point towards distinct, multivariate genetic architectures underlying CT and SA.

The current study utilizes large-scale, imaging genetics datasets to formally model the genetic overlap across brain regions within CT and SA using the Genomic Structural Equation Modeling (Genomic SEM) framework.^10^ We began by performing exploratory and confirmatory factor analyses of the genetic covariances estimated from the ENIGMA CT and SA summary statistics. We then replicate this factor structure in a semi-independent sample from UK Biobank (UKB), showing that the multivariate structure identified in ENIGMA fits the data well for both the left and right hemisphere in UKB. Having established the portability of this factor structure, we characterize the genomic factors at three levels of analysis. First, we examined the boundaries that define these genomic factors by examining relationships with molecular, cellular, and functional topographical maps. Second, we applied Stratified Genomic SEM to identify classes of genes that are enriched at the level of the structural imaging factors. Finally, we examined associations between the brain factors and both general and domain-specific facets of cognitive function and psychiatric disorders. Collectively, our multivariate analyses of structural imaging phenotypes provide key insights into the biological, functional, and clinical relevance of varying levels of structural brain organization.

## Results

### Genomic Factor Analysis

LD-score regression applied to the ENIGMA summary statistics (*N* ≈ 33,992 participants) for the 34 bilateral averages of regional CT and SA revealed high levels of genetic overlap (**Figure 1**). To formally model this pattern of genetic overlap, we began by applying three tests (Kaiser,^11^ acceleration factor, and optimal coordinates^12^) to determine the optimal number of factors that could be used to parsimoniously describe the data. We then fit exploratory factor analyses (EFAs) based on these tests and finally used the EFA results to inform fitting confirmatory factor analytic models (CFAs) within Genomic SEM (additional details provided in **Method; Figure 1** for example path diagram). Genomic SEM results revealed that for both CT and SA a five-factor correlated factors model and bifactor model with five-factors plus a general factor fit the data well, while a single common factor model did not (see Supplementary Tables 1-2 for full model output; Supplementary Table 3 for model fit; Supplementary Figures 1-2 for path diagrams). We note that GWAS summary statistics were not corrected for a global structural metric (e.g., total SA or mean CT). Instead, global (co)variation was accounted for by modeling a latent general factor in the bifactor models. Although the bifactor model ultimately fit the data best for CT and SA, we consider both the bifactor and correlated factors models for all downstream analyses. This decision point in part reflects the observation that bifactor models are generally guaranteed to fit the data better, regardless of the data generating process in the population.^13^ At the same time, we consider the bifactor model informative as brain regions are known to globally covary, and it provides a psychometrically informed comparison point to previous results produced using GWAS summary statistics that controlled for total SA and average CT. By considering both models, we were able to formally quantify variation in the genomic factor structure, and corresponding effects on downstream results, across models that do and do not control for global covariation. In addition, this analytic pipeline has the advantage of avoiding bias due to adjusting for a heritable trait (i.e., global metrics for SA and CT).^14^

**Figure 1.**
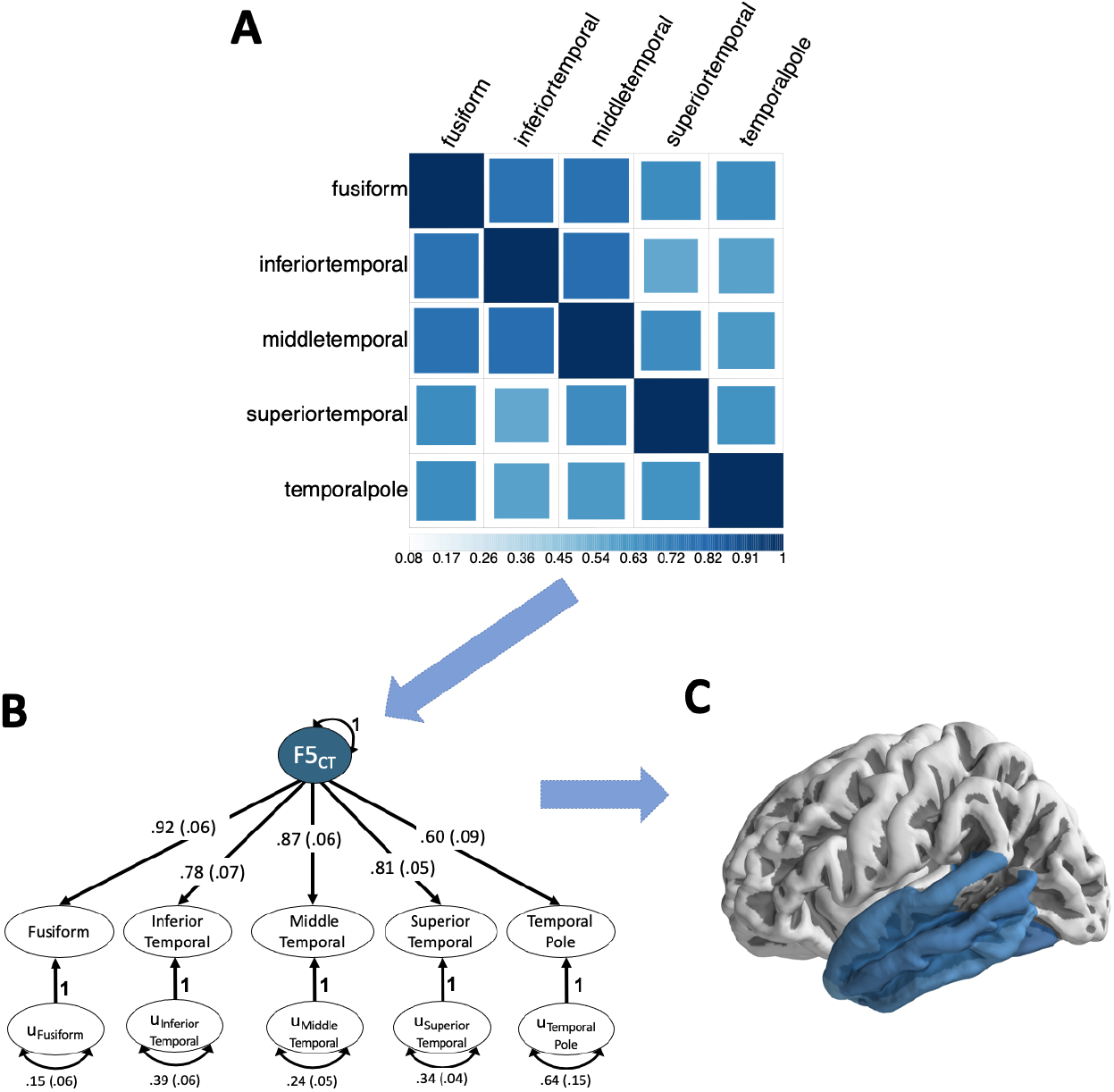
Genomic SEM Schematic. Figure depicts outline of the Genomic SEM factor modeling for a subset of cortical thickness phenotypes that load on a single factor. *Panel A* depicts the genetic correlation matrix estimated using LDSC across the five brain regions that define the fifth factor (F5CT) from the cortical thickness model. These five brain regions were identified as loading on the same factor using the exploratory factor analyses (EFA) detailed in the **Method** section. As would be expected based on the EFA results and can be observed in this genetic correlation matrix, these five traits evinced a particularly high level of genetic overlap. *Panel B* depicts the path diagram for the standardized, confirmatory factor model results produced by Genomic SEM using this same genetic correlation matrix as input. The genetic components of each brain region, the common factor defined by these genetic components, and the residual genetic variance for each brain region are represented as circles to reflect the fact that these are latent (i.e., not directly observed) variables. For model identification purposes, the variance of the genomic brain factor was fixed to 1. We display only this single factor from the CT model as an exemplar, but the broader factor model included the additional four factors, and correlations across these factors, are displayed in **Supplementary Figure 1**. *Panel C* depicts the physical location of these five brain regions that load on a single factor in dark blue shading; we find that all genomic brain factors were defined by contiguous brain regions.

As would be expected, physically proximal brain regions clustered within the same factors (**Figure 2**). For CT, these factors can be described as representing frontoparietal (F1CT), cingulate (F2CT), prefrontal (F3CT), occipital (F4CT), and lateral temporal (F5CT) brain regions. For SA, the first two factors were far more diffuse, representing a combination of frontoparietal and temporal regions (F1SA and F2SA), followed by a third factor (F3SA) defined by occipito-parietal regions, and finally two highly specific factors, one reflecting the rostral and caudal anterior cingulate regions (F4SA), and a fifth factor (F5SA) defined solely by the medial orbitofrontal region. On average, the five factors for each metric were highly genetically correlated (average factor *r*_*g*_ = .80 for CT; average *r*_*g*_ = .82 for SA), with some factors evincing more distinct genetic underpinnings (e.g., *rg =* .66 across the CT cingulate [F2CT] and occipital [F4CT] factors).

In line with these large factor correlations, the average proportion of genetic variation in the individual brain regions explained by the general factor from the bifactor model was 52.1% for CT and 56.7% for SA. In addition, excluding the variation explained by the general factor, there was a 65.8% reduction for CT and 75.9% reduction for SA in the variation explained by the five factors for the bifactor model relative to the correlated factor model. However, 24 of the 34 CT factor loadings and 23 SA loadings on the five factors in the bifactor model remained significant at a Bonferroni corrected threshold for 34 brain regions. Collectively, these results indicate that the pervasive genetic overlap across regional measures of CT and SA are not merely reflective of a single dimension of macroscale organization. Rather, the multivariate genetic architecture of the cerebral cortex can be parsimoniously described using five factors for each metric that explain significant genetic variation across brain regions even when accounting for global covariation.

**Figure 2.**
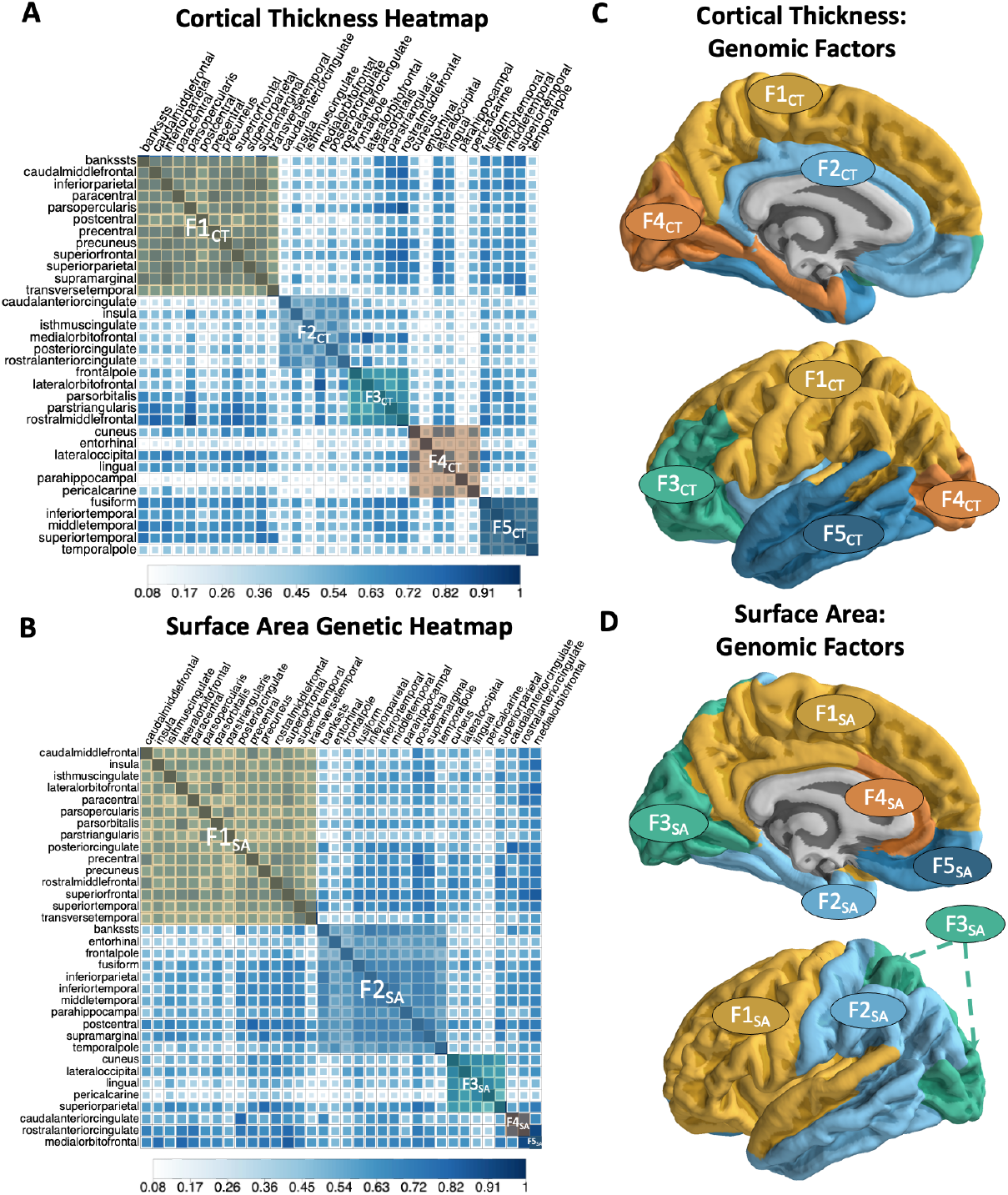
Genomic Factor Analysis of Cerebral Cortex. *Panels A* and *B* depict the genetic heatmaps, as estimated using LD-score regression, for cortical thickness and surface area, respectively. The heatmaps are ordered with respect to the factor model identified in Genomic SEM and the brain regions that define a factor are overlaid with semi-transparent squares with coloring that corresponds to the *Panels C and D. Panels C* and *D* specifically depict the brain regions color coded according to the five genomic factors on which they load. These brain regions are labeled F1 – F5 for comparison to the full set of Genomic SEM results reported in Supplementary Tables 2-3 and depicted in the path diagrams in Supplementary Figures 1-2.

The replicability of these findings was tested using a semi-independent, genetically-informative imaging dataset from UKB (*N* = 26,739; see **Method** for details on sample overlap). We found that the correlated factors and bifactor models fit the data well for both the left and right hemispheres in UKB (Supplementary Table 3 for model fit; Supplementary Tables 4-7 for full model outputs; Supplementary Figures 3-7 for heatmaps), indicating that the identified factor structure is stable and captures a robust, bilaterally symmetrical pattern of genetic covariation.

### Topographical Annotation of Cortical Genomic Factors

We next examined whether spatial organization of the CT and SA factors reflected a biologically- and functionally-relevant partitioning of the cortex. To test these hypotheses, we utilized spin-based methods to compare the factor analytic parcellation to several canonical and meta-analytic maps from the neuroimaging literature (**Method**). First, we asked whether there was statistically non-random overlap in the assignment of regions of interest to CT and SA genomic factors. We find that the spatial organization of both metrics did significantly overlap with one another (*P*spin < 1e-4), suggesting that the genomic factors reflect meaningful boundaries of cortical (co)variation that are partially consistent across these two morphological indices. Moreover, these genomic parcellations appear to capture biological differences of the cortical sheet, as comparisons to a digitized parcellation of von Economo and Koskinas’s^15^ cytoarchitectonic mapping of the cortex (**Figure 3a**) also revealed significant overlap (CT *P*spin = 1.00e-4, SA *P*spin = 3.82E-2, **Figure 3b**,**c**).

**Figure 3.**
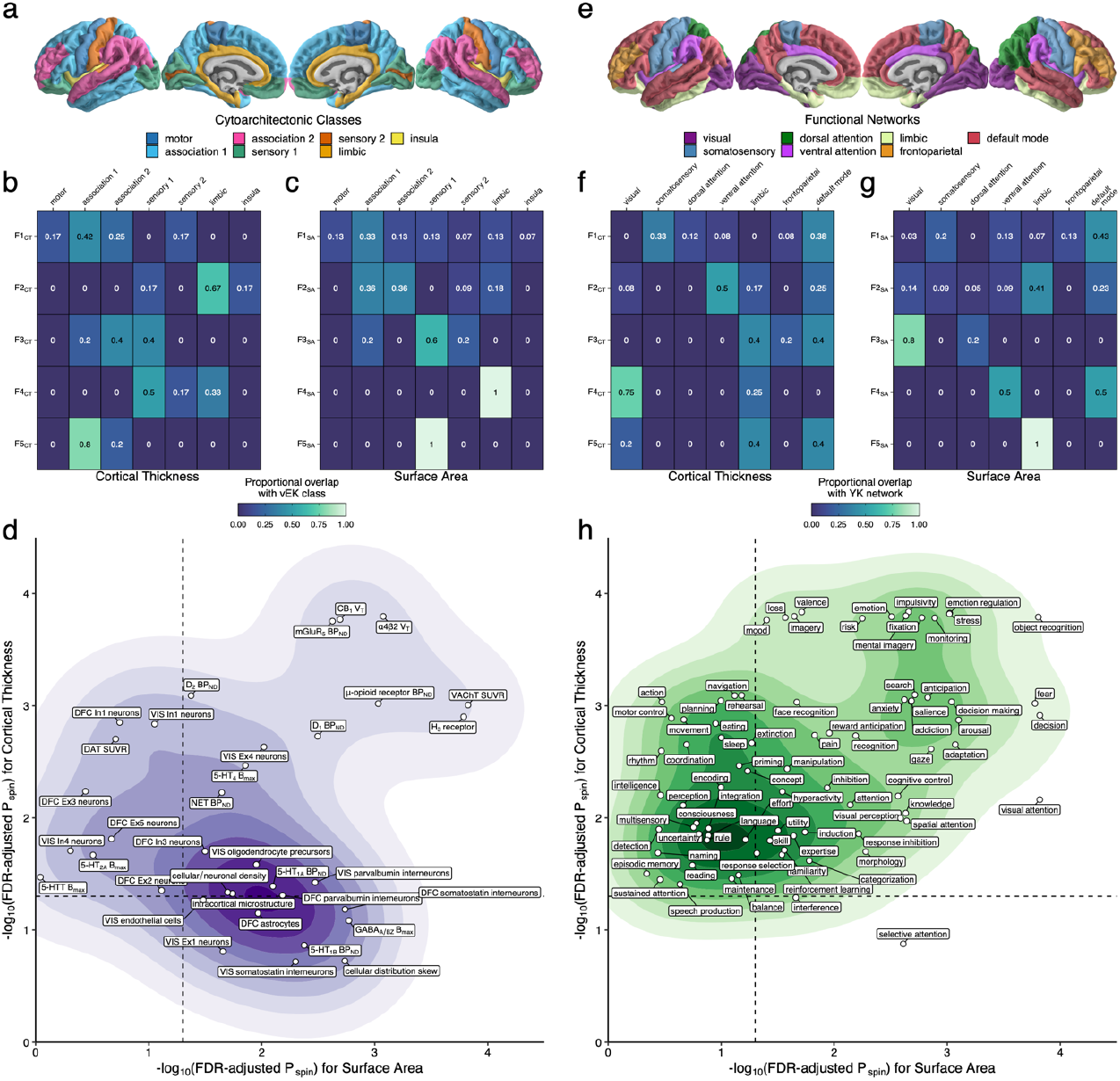
Topographical annotation of genomic cortical thickness and surface area factors. **a**, Map of cytoarchitectonic classes defined by von Economo and Koskinas (vEK)^15^ **b**,**c**, Marginal table of proportional overlap between (**b**) cortical thickness (CT) and (**c**) surface area (SA) and the vEK cytoarchitectonic classes. **d**, Two-dimensional density plot of significant overlap between cortical factors and biologically-derived features of the cortex (from the BigBrain project, Allen Human Brain Atlas, and positron emission tomography meta-analysis; **Method**). The dashed line denotes statistical significance on a -log10 scale after correcting for false discovery rate (FDR). **e**, Map of canonical resting-state functional connectivity networks defined by Yeo and Krienen (YK).^16^ **f**,**g**, Marginal table of proportional overlap between (**f**) CT and (**g**) SA and the YK networks. **h**, Two-dimensional density plot of significant overlap between cortical factors and patterns of functional activation associated with psychological processes (from Neurosynth; **Method**). The dashed line denotes statistical significance on a - log10 scale after correcting for FDR.

Consistent with the notion that these genomic factors differentiate broad areas of the cortex by their biological underpinnings, further topographical annotation identified myriad aspects of intracortical microstructure, laminar differentiation, cellular/neuronal density, neurotransmitter receptor density, and cell type-specific transcriptional signatures that significantly varied as a function of the CT and SA factors (**Figure 3d**, Supplementary Figures 7-8, Supplementary Table 9). Generally, results suggest that the SA factors better captured interregional variation of these biological features relative to the CT factors (**Figure 3d**). While similar effects were seen for some neurobiological features (e.g., CB1, D1, and μ-opioid receptor densities), CT- and SA-specific effects were also observed, such as In1 inhibitory neuron signatures that were more associated with CT and and somatostatin interneuron transcriptional signatures more associated with SA.

Given these findings of molecular and cellular differences, we next asked how the genomic parcellations were organized relative to the functional topography of the cortical sheet. We first compared the spatial organization of the genomic factors to that of the seven canonical functional networks defined by Yeo and Krienen^16^ (**Figure 3e**), which revealed significant overlap between these maps (CT *P*spin = 1.20E-3, SA *P*spin =7.70E-3, **Figure 3f**,**g**). To obtain finer-grained insights into how the genomic factors might relate to aspects of brain function, we then compared our factor analytic maps to meta-analytic maps of functional activation for 123 cognitive processes from Neurosynth (**Method**). As observed with the biologically-focused analyses above, we found that many patterns of functional activation associated with cognitive processes, emotion regulation, reward learning, decision-making, and visual processing were significantly different across the genomic factors for both CT and SA (**Figure 3h**, Supplementary Figures 9-10, Supplementary Table 9). However, in contrast to the more biologically-derived maps, these results revealed that CT factors better captured interregional variation of psychological and cognitive features relative to the SA factors, with specific effects for perception, locomotion, and language.

### Stratified Genomic SEM

Stratified Genomic SEM is a multivariate analogue of Stratified LDSC^17^ that can be used to determine whether a given functional annotation is enriched for any model parameter within Genomic SEM. Functional annotation is used here to denote a set of genetic variants that are grouped according to shared characteristics. For the current analyses, we specifically utilize 168 functional annotations reflecting genes expressed in different brain regions, evolutionarily conserved regions, tissue types, histone marks, neuronal cell types, and protein truncating variant intolerant (PI) genes (**Method** for further details). A functional annotation is then determined to be enriched if the proportion of genetic variance explained by the annotation is greater than the proportional size of the annotation. Consequently, if a small annotation defined by relatively few genetic variants explains an outsized proportion of genetic variation, then that annotation will be identified as highly enriched. Multivariate functional analyses thereby allow for distilling both highly polygenic and pleiotropic signals into biologically meaningful, constituent parts. To this end, we specifically examined enrichment of the factor variances in the correlated factors and bifactor models.

For CT, 9 annotations were found to be significantly enriched for at least one of the factors from the correlated factors model (Supplementary Table 10). This included: significant enrichment of the H3K27ac cingulate gyrus histone mark, endothelial cells, and PI genes for F1CT; oligodendrocytes and the H3K4me1 cingulate gryus histone mark for F2CT; and the middle hippocampus H3K27ac histone mark and PI genes for F3CT. In the bifactor model, there were 2 significant annotations for the general factor: H3K4me1 union and the cingulate gyrus H3K27ac histone mark.

We identified 13 significant annotations for SA in the correlated factors model (**Figure 4**; Supplementary Table 11). This included significant enrichment of endothelial cells and the H3K27ac histone mark within the anterior caudate for F1SA; the intersection of microglia and PI genes for F2SA; and the H3K4me3 histone mark in the dorsolateral prefrontal cortex and H3K27ac histone mark in the angular gyrus for F3SA. There were 14 significant annotations in the bifactor model for SA. This included 7 annotations that were originally significant for one of the correlated factors, but were identified as enriched for the general factor in the bifactor model. Consistent with the observation that the PI genes and Super Enhancer annotations were significant for ≥ 4 factors in the correlated factors model, we find that these are the two most enriched annotations for the general factor in the bifactor model. This was followed by the H3K4me3 histone mark in the fetal female brain and astrocytic transporters. No annotations that were significant in the context of the correlated factors model remained significant in the bifactor model for either CT or SA.

**Figure 4.**
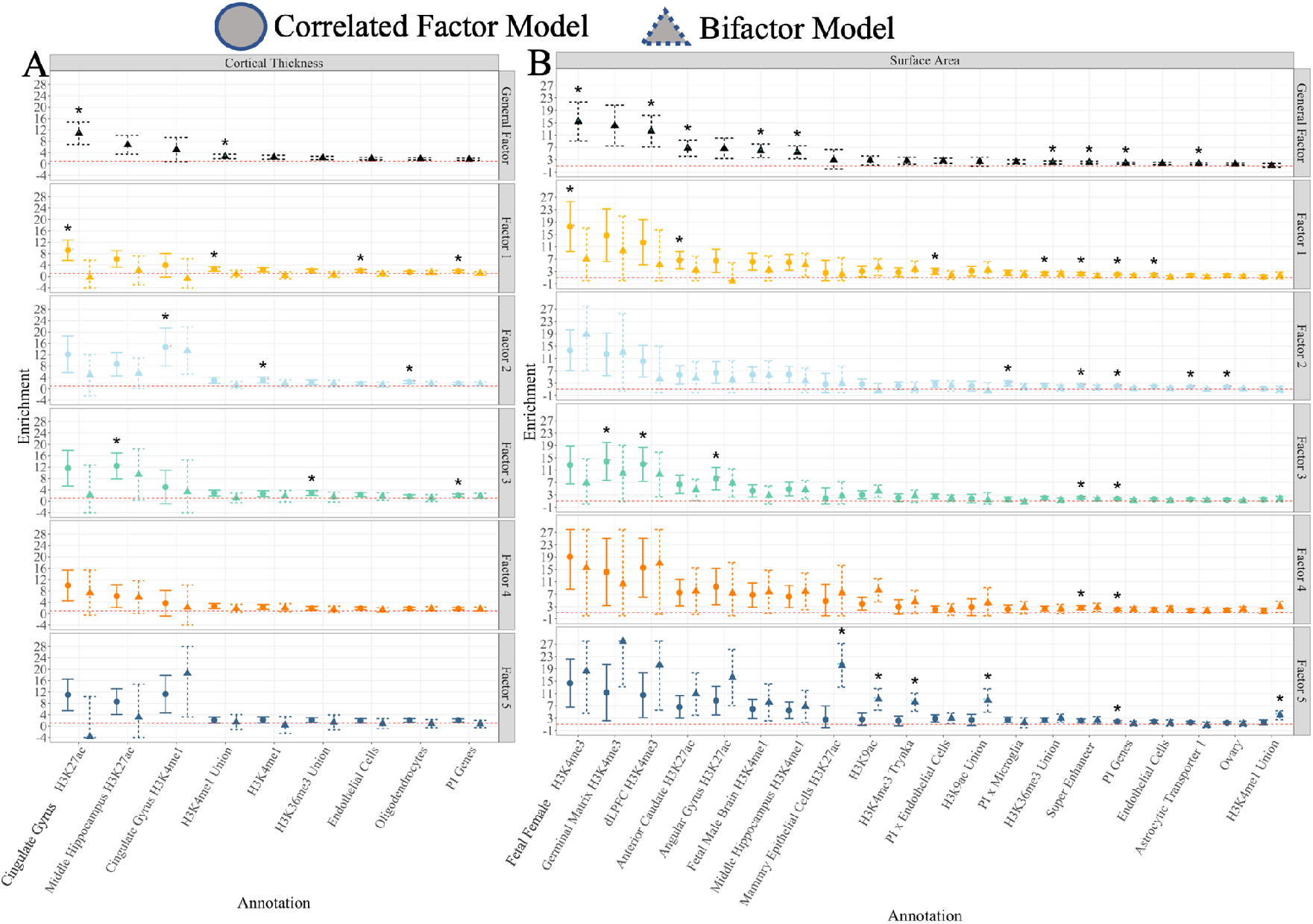
Stratified Genomic SEM Results. Figure depicts the 9 functional annotations for Cortical Thickness (*Panel* ***A***) and 20 annotations for Surface Area (*Panel* ***B***) that were significantly enriched at a Bonferroni corrected threshold for at least one of the factors for either the correlated factor or bifactor model results. Error bars reflect 95% confidence intervals and are capped at the y-axis limits for visualization purposes. Significant annotations are depicted with a *. For Factors 1-5, results depicted in circles with solid error bars reflect results from the correlated factors model and triangles with dashed line error bars results from the bifactor model. Results for the general factor at the top of each panel are for the common factor defined by all 34 brain regions in the context of the bifactor model. The horizontal red dashed line at 1 reflects the null of no enrichment.

### Genetic Overlap with Cognitive Function

We examined genetic overlap between the brain-based factors and a *g*-factor estimated from UKB GWAS summary statistics for seven cognitive traits: trail-making tests-B, tower rearranging, verbal numerical reasoning (VNR), symbol digit substitution, memory pairs-matching test, matrix pattern recognition, and reaction time (RT). The shared genetic covariance across these seven cognitive indicators was modeled using the same common factor model for genetic *g* identified by de la Fuente and colleageus,^18^ and genetic correlations were estimated for *g* and the brain-based factors for the separate CT and SA models. We first sought to test whether the degree of genetic overlap across *g* and brain morphology also varied by factor. For both CT and SA, we found robust evidence that the genetic correlations between *g* and the brain factors could not be constrained to be equal (*p* < .05 for both χ^2^ difference tests, **Method**). This suggests that there is significant variation across the cortical sheet in the degree of genetic overlap between intelligence and brain structure.

We next turned our attention to both global and localized patterns of genetic overlap between the brain factors and *g*, allowing for residual covariances between individual brain regions and cognitive tests where indicated (**Method**). As each family of tests for CT and SA with *g* consisted of 204 possible associations (i.e., 34 brain regions × 7 cognitive tests), we employ a Bonferroni correction of .05/204 (*p* < 2.10E-4). Results for SA revealed significant genetic correlations between all five factors and the *g*-factor in the context of the correlated factors model (average *r*_*g*_ = .22; range = .16-.25; Supplementary Table 12). Bifactor model results revealed a significant genetic correlation between the general SA and *g-*factor (*r*_*g*_ = .24, *SE* = .04, *p* = 2.34E-11), while none of the correlations between *g* and the five residual factors remained significant. There were no significant factor correlations for CT in the correlated factors or bifactor model after correcting for multiple comparisons. However, we observe significant overlap across the residual genetic variance for VNR (also termed fluid intelligence in UKB) and the precentral region for the correlated factors model (partial *r*_*g*_ = .31, *p* = 1.02E-4). These results indicate that the genetic overlap between SA and cognitive function can be conceptualized as operating through largely general pathways shared across brain regions and different classes of cognitive tasks. Conversely, results for CT reflected far more circumscribed patterns of overlap specific to individual cognitive tasks/brain regions.

### Genetic Overlap with Psychiatric Traits

To examine the multivariate system of genetic relationship with psychiatric traits, we employed the same four-factor correlated factors model of 11 major disorders identified in Grotzinger al. (2020).^19^ This model consists of a Compulsive disorders factor defined by anorexia nervosa,^20^ obsessive compulsive disorder (OCD),^21^ and Tourette’s syndrome,^22^ a Psychotic disorders factor defined by schizophrenia^23^ and bipolar disorder,^24^ a Neurodevelopmental disorders factor defined by autism spectrum disorder (ASD),^25^ attention deficit hyperactivity disorder (ADHD),^26^ and post-traumatic stress disorder (PTSD),^27^ and an Internalizing disorders factor defined by major depressive disorder (MDD),^28^ anxiety disorders,^29^ and PTSD^27^ (Supplementary Table 13 for factor loadings from psychiatric measurement model). In addition, alcohol use disorder^30^ loaded on the Psychotic, Neurodevelopmental, and Internalizing factors. As with the *g*-factor analyses, we tested whether the degree of genetic overlap between psychopathology and brain morphology varied across the cortex. For CT, the constrained model with invariant genetic correlations per psychiatric factor would not converge, suggesting it is not an appropriate model. Similarly, for SA, we found robust evidence that the genetic correlations between the psychiatric factor and the brain factors could not be constrained to be equal (*p* < .05, **Method**), indicating that there is significant variation across the cortex in the degree of genetic overlap with the psychiatric factors.

We then estimated genetic correlations between all psychiatric and brain factors, and added residual covariances between individual disorders and brain regions when indicated. We employ a Bonferroni corrected significance threshold of *p* < 1.34E-4 (**Method**). Results for CT revealed no significant correlations between any of the brain and psychiatric factors in either the correlated factors or bifactor model (Supplementary Table 14). For SA, a significant, negative genetic correlation was observed between the Neurodevelopmental disorders factor and F2SA, defined by temporal and frontal regions (*r*_*g*_ = -.22, *SE* = .05, *p* = 2.43E-5). While this correlation was not significant in the bifactor model, it is of note that the Neurodevelopmental factor, relative to the other psychiatric factors, evinced a pattern of generally stronger, negative associations with the other brain factors. Consistent with this observation, the Neurodevelopmental factor displayed the strongest genetic correlation with the general SA factor in the bifactor model (*r*_*g*_ = -.17, *SE* = .05, *p* = 1.19E-3), though this was not significant at a Bonferroni corrected threshold. Four nominally significant residual relationships were also identified between ASD and specific brain regions, the strongest of which was with the rostral anterior cingulate for both the correlated factor (partial *r*_*g*_ = .180, *SE* = .051, *p* = 4.06E-4) and bifactor model (partial *r*_*g*_ = .184, *SE* = .051, *p* = 3.23E-4).

## Discussion

The current study examined the multivariate genomic architecture of CT and SA at various levels of analyses using the largest available imaging genomic datasets. At the genome-wide level, we find that ENIGMA summary statistics for 34 physically proximal brain regions can be grouped across five, correlated genomic factors. In addition, we observe that this factor structure fits the data for the left and right hemisphere in UK Biobank, indicating a bilaterally symmetrical structure, and that these five factors continue to explain significant genetic variation in the brain indicators when pulling out shared global variation via a general factor in a bifactor model. It is of note that the multivariate architectures with respect to the make-up of the genomic factors were distinct across CT and SA. This is in line with prevailing developmental accounts of these structures, including the radial unit hypothesis that argues for distinct developmental trajectories categorized by neurogenetic division of neural progenitor cells for CT as compared to propagation of these cells for SA.^31^ Indeed, previous studies have shown that CT and SA are characterized by differing developmental trajectories,^32,33^ and the family-based literature corroborates the finding that genetic variation in these two metrics are distinct.^7,8^

To understand these novel genomic parcellations of the cortex, we used recently developed surface-based methods to place our findings in the broader context of the neuroimaging literature. This topographical annotation allowed us to evaluate the spatial organization of our CT and SA factors relative to two canonical parcellations of the cortex: the cytoarchitectonic classes defined by von Economo and Koskinas^15^ and the functional networks derived by Yeo and Krienen.^16^ For both canonical parcellations, we found significant patterns of overlap between classifications of cortical regions, suggesting the organization of the CT and SA factors reflected a biologically- and functionally-relevant partitioning of the cortex. Further annotation revealed relationships with myriad quantitative features of the cortex, such as neurotransmitter receptor densities, cell-type transcriptional signatures, and patterns of functional activation measured by fMRI. These results provide an anatomical context for our factor analytic findings and lend insight into what underlies the differences between CT and SA factors. For example, while the SA factors were perhaps more broadly related to biologically-derived features of the cortex (e.g., neuronal density, neurotransmitter receptor densities), the CT factors were more strongly related to functional activation underlying a variety of psychological processes. Interestingly, the spatial organization of both CT and SA factors captured functional boundaries of the cortex underlying emotion regulation, reward learning, decision-making, and visual processing.

At the functional genomic level, we observe a number of annotations that were significantly enriched for CT and SA. In some cases, annotations enriched at the factor level mapped onto the regions that defined the factor. For example, we observe that the cingulate factor (F2CT) for CT was enriched for the tissue specific expression of the H3K4me1 histone mark in the cingulate gyrus. We find also that astrocytic transporters and, consistent with their generalized biological roles, the PI genes and Super Enhancer annotations are broadly relevant for SA, with significant enrichment identified across factors in the correlated factors model and for the general factor in the bifactor model. Notably, we do not observe enrichment for these annotations for CT, indicating that these classes of genes are specifically relevant to SA. It can also be informative to consider how the patterns of enrichment identified here diverge from enrichment identified by ENIGMA for their global metrics of CT and SA.^9^ For example, endothelial cells were highly enriched for global CT in ENIGMA, but were found to be uniquely enriched for the factor defined by central regions in the current analyses (F1CT). Similarly, microglia were significant for global SA in ENIGMA, but only for F2SA in the current analyses. These findings tentatively indicate that prior enrichment findings for the global signal may be collapsing across patterns of enrichment unique to subclusters of contiguous brain regions.

Consistent with prior literature,^3–6^ we observe that genetic variation in SA is broadly relevant for a range of cognitive functions, as indexed by a genetic *g*-factor defined by seven cognitive outcomes. More specifically, we observe significant genetic correlations between *g* and the five factors from the SA correlated factors model and the general factor from the bifactor model. Results were far more limited for CT, but included a significant, negative genetic correlation between the precentral region and verbal numerical reasoning, a measure of fluid intelligence. While this latter result should be reexamined as sample sizes continue to grow, it is consistent with prior findings from a semi-independent sample.^34^

With respect to genetic overlap with psychiatric factors, we found that the Neurodevelopmental disorders factor (indexed predominantly by PTSD, ADHD, and ASD) was negatively associated with F2SA (defined by temporal and frontal regions). While this correlation did not remain significant in the bifactor model, the Neurodevelopmental factor displayed a generally pervasive pattern of negative associations with SA that, given the stringent Bonferroni correction applied here, should be reevaluated as larger samples become available. We also observe positive, residual genetic overlap across ASD and four brain regions, the strongest of which was observed for the rostral anterior cingulate region. Perturbations in the cingulate region more broadly have also been observed for prior phenotypic imaging and ASD studies.^35,36^ The current findings are then informative for interpreting this prior literature given high levels of genetic and phenotypic overlap across psychiatric and structural imaging phenotypes. That is, the multivariate genomic analyses performed here indicate that ASD-cingulate associations can be considered as highly specific to these two phenotypes, as opposed to an indirect proxy for far more generalized pathways across highly correlated psychiatric and structural outcomes.

In contrast to prior findings, we observe no significant genetic relationships between CT and psychiatric traits after correcting for multiple comparisons. We view this null finding as informative in its own right with respect to where we “point the telescope” next in psychiatric genetics. That is to say, the current analyses reflect a well-powered, psychometrically informed approach to examining genetic overlap across CT and a broad range of clinical presentations. These null results may then help to narrow the search space for psychiatric risk factors/correlates and should be considered when interpreting prior results linking CT to psychiatric disorders. Prior findings may then reflect false positives, associations that operate through largely environmental pathways, associations between variation in CT unique to clinical samples that is not captured using a general population imaging sample, associations that are developmentally bound, or some combination of the four.

The current analyses have a number of limitations that should be noted. First, we highlight that these results are restricted to participants of European ancestry due only to the availability of sufficiently well-powered GWAS data for this ancestral group coupled with the requirement of LD-score regression to produce estimates within a single ancestral population due to differences in LD structure across groups. It will be of the utmost importance both with respect to scientific value and representation that future analyses build on the expanding, genetically informed datasets for different populations. While we were able to evaluate the fit of the factor structure in both ENIGMA and UK Biobank, these were not entirely independent samples. In addition, it is of note that ENIGMA reflects an age heterogeneous sample, and findings should be interpreted in this light. These models should continue to be evaluated in external, genetically informed imaging datasets for specific developmental windows. For Stratified Genomic SEM analyses, we utilized the zero-order stratified genetic covariance matrices that do not control for overlap with other annotations for estimation of enrichment. This decision point reflects the power needed to utilize the *τ* matrices that do control for annotation overlap (**Method**), but as GWAS sample sizes continue to grow, future work can examine the robustness of these results.

Substantially overlapping genetic signal within CT and SA brain regions necessitates statistical tool that allow for examining the multivariate system of relationships across these phenotypes. To this end, we examined the factor structure and its correlates at the genome-wide level, and went on to better characterize these factors by performing topographical annotation and estimating functional enrichment. In line with prior studies that utilize the global metrics of CT and SA, we find that a general factor explains significant genetic variation and captures patterns of enrichment shared across the 34 brain regions. At the same time, we observe both that the five genomic factors reflect biologically and functionally relevant partitionings of the cortex and that certain associations with external correlates and patterns of enrichment are unique to either the factors or specific brain regions. As all data used here is publicly available, we present an accessible, novel framework for studying the multivariate genomic architecture of the cerebral cortex. We propose that future studies use this approach to examine other research questions relevant to this highly studied set of imaging outcomes. Collectively, these findings point towards the utility and need to simultaneously model the different levels of genetic risk sharing, from the most general level across all the cortex, to the specific subclusters indexed by the factors, and down to the variation unique to a single brain region.

## Method

### Quality Control Procedures

Quality control filters for estimating the genetic covariance and sampling covariance matrices followed the defaults in the Genomic SEM implement of LDSC. These filters included restricting to SNPs present in HapMap3 with a minor allele frequency > 1% and information scores (INFO) > .9. The LD scores used for LDSC were calculated using the European subsample of the 1000 Genomes phase 3 project; the scores excluded the MHC region due to the high degree of LD outliers which is known to unduly influence estimates. We note also that when calculating the liability scale heritability for the psychiatric traits we used the sum of effective sample sizes, and a sample prevalence of .5 to reflect the fact that the corrected sample size already accounts for sample ascertainment; we have shown that this produces a more accurate estimate of heritability for binary traits as it more appropriately accounts for ascertainment differences across cohorts contributing to GWAS meta-analysis.^37^ For comparative purposes, population prevalences for the liability conversion were chosen to reflect those used from the corresponding univariate GWAS publication and are reported in Supplementary Table 11.

### Genomic Factor Analysis: ENIGMA

We refer the reader to the original ENIGMA^9^ publication for details about how the univariate GWAS were performed. We note briefly here that ENIGMA^9^ used the Desikan-Killiany^38^ atlas segmentations to define the 34 brain regions and that SA and CT were measured using T1-weighted magnetic resonance imaging scans. We used the summary statistics that are made publicly available that apply genomic control. We note also that all analyses presented here utilize the GWAS summary statistics that were not corrected for global volume as this allowed for explicitly modeling the shared genetic variation across the 34 regions in the context of a bifactor model. Factor analysis of ENIGMA GWAS summary statistics proceeded in five primary steps. First, standard quality control filters (see QC section below) were applied to the GWAS summary statistics using the *munge* function in Genomic SEM. Second, LD-score regression^39,40^ was applied to the ENIGMA GWAS summary statistics, in combination with the 1000 Genomes Phase 3 European LD scores that excluded the MHC region, to produce a genetic heatmap across the 34 CT and SA brain regions. As certain SNPs were not present across all cohorts that comprise the ENIGMA consortium, the SNP-specific participant sample sizes were used for LDSC estimation. Third, the Kaiser,^11^ acceleration factor, and optimal coordinates^12^ rules were applied to these genetic correlation matrices in order to collectively determine the number of genomic factors that could be used to parsimoniously represent the data. For both SA and CT, these results pointed towards five factors according to the Kaiser and optimal coordinates tests, and a single, common factor according to the acceleration factor test. Fourth, exploratory factor analyses (EFAs) were conducted using the promax (i.e., correlated factor) rotation in the *factanal* R package.

Finally, we fit confirmatory factor models in Genomic SEM and evaluated these models using standard metrics of model fit.^10^ More specifically, comparative fit index (CFI) values above .9^41^ and standardized root mean squared residual (SRMR) values less than .10 were considered indicative of acceptable model fit. We also report the Akaike Information Criteria (AIC)^42^ a fit index that balances overall model fit with the number of estimated parameters (i.e., parsimony), with lower values indicating better fit. We fit three primary confirmatory models in Genomic SEM. The first was a common factor model that was used to determine whether, consistent with the acceleration factor test, a single factor was sufficient for describing the data. The second was a five factor correlated factors model specified based on the five-factor EFA results. More specifically, individual brain regions were assigned to a factor when their standardized loading was > .5, or if the brain region did not achieve a loading of .5 for any factor assigning the region to the factor with the largest standardized loading.^*1*^ This was with the one exception that the medial orbitofrontal region was the one indicator that showed evidence of cross-loadings for both CT and SA, with standardized loadings > .5 for both the second and third factor in the CT model and the first and fifth factor in the SA model. However, including this cross-loading in the confirmatory model produced worse model fit for CT (with cross loading: AIC = 68157.97, CFI = .924, SRMR = .070; without cross loading: AIC = 53570.8; CFI = .941; SRMR = .070); it also included the only negative factor loadings for CT, and caused model convergence issues for SA. This cross-loading was consequently removed for both structural metrics with the medial orbitofrontal factor specified to load on the factor with the highest EFA loading.

The third type of model we fit was a bifactor model, which consisted of a general factor that captures shared variation across the 34 brain regions and five, residual factors (defined by the same brain regions from the correlated factors model) that model covariation not accounted for by the general factor. As the general factor defined by all indicators within a bifactor model is conceptually posited to account for the covariation across the remaining factors, the five residual factors were all specified to be orthogonal (i.e., factor correlations fixed to 0). For all models, we used unit variance identification such that the factor variances were fixed to 1. For the SA models, the fourth factor was defined by two brain regions (caudal anterior cingulate and rostral anterior cingulate) and the fifth factor was defined only by the medial orbitofrontal brain region. To ensure that the SA models were locally identified, the factor loadings were then constrained to equality for the fourth factor and the residual variance of the medial orbitofrontal region that solely defined the fifth factor was fixed to 0.

Given the pervasive levels of genetic overlap across the 34 brain regions, the generally parsimonious representation of the data using five factors, and the stringent threshold of assigning brain regions to factors using standardized loadings of .5 or greater, we went on to iteratively add residual covariances across pairs of brain regions for the correlated factors model. This was done by obtaining the residual covariance matrix—calculated as the difference between the model-implied genetic covariance and observed genetic covariance matrix—and adding the residual covariances one at a time until they no longer reached a significance threshold of *p* < .01. This procedure resulted in adding eight residual covariances for CT and seven residual covariances for SA. We confirmed that these residual covariances improved model fit for both the correlated factors and bifactor model for CT and SA (Supplementary Table 4).

Although the bifactor model with residual covariances ultimately fit the data best for CT and SA, we consider both the bifactor and correlated factors models with residual covariances for all downstream analyses. This decision point in part reflects the observation that bifactor models are generally guaranteed to fit the data better, regardless of the data generating process in the population.^13^ At the same time, we consider the bifactor model informative as brain regions are known to globally covary, and it provides a psychometrically informed comparison point to previous results produced using GWAS summary statistics that controlled for total SA and average CT. In summary, we find for both CT and SA that a bifactor model and five-factor correlated factors meet or exceed field standard metrics for providing acceptable fit to the data and are both considered theoretically informative for downstream analyses, including replication in UK Biobank.

### Genomic Factor Analysis: UK Biobank

The UK Biobank (UKB; http://www.ukbiobank.ac.uk) is a large population-based cohort study that recruited approximately 500,000 volunteers between 2006–2010 across the UK. A subset of participants underwent brain MRI scans since 2014. UKB received ethical approval from the North West Centre Research Ethics Committee (REC number 11/NW/0382). The current analyses were conducted under the approved UKB application 32568.

Raw brain imaging data were processed through an automated image processing pipeline^43^ by the UKB imaging team to create a wide range of image-derived phenotypes (IDPs) (https://www.fmrib.ox.ac.uk/ukbiobank/fbp). Details of the MRI protocol and processing are publicly available.^43,44^ Here, we focused on CT and SA measures of the 34 brain regions defined by the Desikan-Killiany atlas.^38^ The temporal pole was excluded from analysis, as part of the temporal lobe is difficult to segment and lead to a large amount of missing data for the region.

Our GWAS analyses began with an initial pool of 40,733 individuals who had MRI data at the time of our analysis. We retained 27,220 individuals of European ancestry whose genetic data passed the quality control. We then removed individuals with incomplete data on necessary covariates (see below), which yielded a final sample size of 26,739. The GWAS analysis was performed using PLINK (https://www.cog-genomics.org/plink/2.0/assoc). Specifically, we used the linear regression model, adjusting for age, sex, X/Y/Z/T position of head and the radio-frequency receive coil in the scanner, UKB imaging acquisition center, mean resting-state and task-based functional MRI head motion, volumetric scaling factor, T1 density, genotyping chip and the top 40 principal components of the genetic data as covariates.

The ENIGMA imaging sample utilized in the primary analyses also includes an earlier release of the UKB imaging data for *N* = 10,083 participants.^9^ As ENIGMA provides GWAS summary statistics, as opposed to individual genotypes, we were unable to curate an entirely independent replication sample within UKB. While the 10,083 UKB participants included in ENIGMA will be encompassed by the total, updated imaging sample of 40,733 UKB participants, at the same time it is highly unlikely that all 10,083 UKB participants in ENIGMA were included in our final sample of 27,739 UKB individuals given different analytic and QC pipelines. We also quantify the level of shared information across our UKB summary statistics and the ENIGMA summary statistics using the bivariate LDSC intercept. The bivariate (i.e., cross-trait) LDSC intercept is estimated directly from the GWAS summary data, and for two traits (1 and 2) is expressed as:

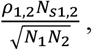

where *ρ*_1,2_ is the phenotypic correlation, *N*_*S*1,2_ is the sample overlap, and *N*_*1*_ and *N*_*2*_ reflect the total sample size for traits 1 and 2, respectively. The bivariate LDSC intercept then reflects the phenotypic correlation weighed by proportional sample overlap, thereby providing a quantitative index of the sampling dependence across the ENIGMA summary statistics and our UKB summary statistics for 27,739 individuals. We specifically estimated the bivariate intercept for global CT and SA metrics for ENIGMA with the global metrics for the left and right hemispheres in UKB. For CT, the bivariate intercept was .178 across ENIGMA and both global metrics of the left and right hemisphere in UKB. For SA, the bivariate intercept was .262 across ENIGMA and the left and right global metrics in UKB. As expected, this indicates that our updated UKB summary statistics reflect a largely independent replication cohort relative to the primary ENIGMA analyses.

### Topographical Annotation

In order to better understand the spatial organization of our genomic brain factors, we sought to characterize their relationships with established canonical and meta-analytic maps from the neuroimaging literature. The specific test employed was dependent on the nature of the comparison (i.e., comparing two categorical maps required a different test than comparing one categorical map and one continuous map), as described in the following sections. However, regardless of the specific test used in a given comparison, a “spin test” approach was used to assess the statistical significance of the observed spatial correspondence. We refer the reader to previous publications for a detailed description of this method, but it critically allows for two neuroimaging maps to be compared while accounting for spatial contiguity and hemispheric symmetry of the cortex. Here, we used the spin test approach to generate an empirical null distribution of 10,000 spatially-permuted test statistics.

To examine the spatial correspondence between our genomic factors of brain morphology and other categorical neuroimaging maps (e.g., the cytoarchitectural classes defined by von Economo and Koskinas,^15,45^ the functional intrinsic connectivity networks derived from resting-state functional magnetic resonance imaging [fMRI] by Yeo and Krienen^16,46^), we first assigned labels from each parcellation to corresponding Desikan-Killiany regions on the basis of maximal overlap. We then performed a Fisher’s exact test to evaluate the degree of dependence between the two categorical maps of interest. The observed *p* value from this test was subsequently compared against an empirical null distribution generated via the spin test approach described above, yielding a final *P*spin value for interpretation.

We also compared the spatial relationships between our genomic factors and numerous features that varied continuously across the cortex, which can be generally described as being derived from either biological/physiological or cognitive/psychological data sources. The former is a collection of cortical maps derived from the BigBrain project (intracortical microstructure, laminar differentiation, cellular/neuronal density)^47–49^ a recent meta-analysis of neurotransmitter receptor and transporter densities measured with positron emission tomography^50^ and the Allen Human Brain Atlas (cell-type specific transcriptional signatures)^51^ while the latter is a collection of cortical association maps obtained from Neurosynth^52^ an online platform for automated meta-analysis of more than 15,000 published fMRI studies. Here, we used a subset of 123 probabilistic association maps that correspond to cognitive and psychological processes described in the Cognitive Atlas^53^ as described in a previous study using similar methods.^50^ For these continuous maps, we fit an analysis of variance model and computed an omnibus *F* statistic for each comparison. As done for the categorical comparisons, *P*spin values were subsequently calculated and adjusted for false discovery rate.

### Stratified Genomic SEM

Stratified Genomic SEM reflects a multi-step process that ends in the ability to estimate enrichment within functional annotations for any parameter of interest (e.g., factor variances) within the genomic model. Functional annotations reflect a subset of genetic variants that are categorized using collateral gene expression data, such as single-cell RNA sequencing, with respect to some shared characteristic, including upward or downward expression within specific tissue types, histone marks, or brain regions. A functional annotation is enriched when the proportion of genetic variation explained by the variants within that annotation is greater than the proportional size of the annotation. The first step in Stratified Genomic SEM is to estimate genetic covariance matrices stratified across a chosen set of annotations. This is achieved by estimating the multivariable version of Stratified LDSC^17^ More specifically, stratified genetic covariance within a functional annotation that controls for overlap with other annotations is estimated as:

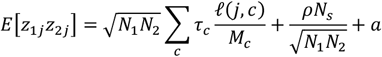

where *N*_*i*_ is the sample size for study *i, c* is a specific genomic annotation, *M*_*c*_ is the number of SNPs in the annotation, *τ*_*c*_ is the coheritability within annotation *c* controlling for overlap with other annotations, *N*_*s*_ is the sample overlap across the two GWAS studies, *a* is a constant term across annotations that captures unmeasured confounding (e.g., shared population stratification), and *ρ* is the phenotypic correlation within overlapping participants. Stratified heritability estimates are produced using the same general formula reduced to the univariate S-LDSC model,^15^ where the expectation across two *z*_*j*_ statistics for the same trait is given as 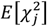.

For the current application, the *τ*_*c*_ values are converted to their zero-order form that does not control for overlap with other annotations. The zero-order stratified values are useful as the estimates are not directly contingent on the other annotations included in the model and produce more stable estimates at moderate GWAS sample sizes, such as those observed here. This comes with the caveat that enriched signal can, in part, reflect signal shared with other annotations and this limitation should be kept in mind when interpreting results. The *τ*_*c*_ estimates are converted to zero-order values (*ζ*_*t*_) for a target annotation *t* by taking the sum of weighted *τ*_*c*_ values given as:

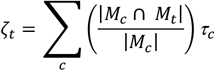

where, as in the bivariate S-LDSC equation described above, |*M*_*c*_| is the total number of SNPs in annotation *c*, and |*M*_*c*_ ∩ *M*_*t*_| is the number of SNPs in both annotation *c* and *t*. Putting these pieces together, 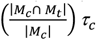 then reflects the stratified (co)heritability estimate in annotation *c* weighted by the proportion of annotation *c* SNPs that are shared with target annotation *t*. The summation of these estimates then produces the zero-order estimates used to populate the zero-order, stratified genetic covariance matrix across the pairwise combinations of included phenotypes.

Each stratified, zero-order genetic covariance matrix is also paired with a stratified, zero-order sampling covariance matrices. The diagonal of the sampling covariance matrix contains squared standard errors of stratified heritability and covariance estimates. The off-diagonals reflect the sampling covariances that capture dependencies among estimation errors that can arise in cases such as participant sample overlap. Both the diagonals and off-diagonals are estimated using a multivariate block jackknife and taken together allow for producing unbiased standard errors in the context of Stratified Genomic SEM. The stratified, zero-order genetic covariance and sampling covariance matrices were specifically estimated using the *s_ldsc* function within the *GenomicSEM* R package.

When the parameter being examined for enrichment reflects pleiotropic effects captured by the factor variances, these stratified matrices are used as input to Genomic SEM wherein the factor loadings are fixed from the model estimated using the genome-wide annotation that includes all SNPs and the factor and residual indicator variances are freely estimated. The factor variances estimated within an annotation then reflect the proportion of pleiotropic genetic variation captured by a given annotation. This estimate and its standard error are subsequently divided by the proportional size of the annotation to produce estimates of multivariate enrichment.

The current project utilized functional annotations from various sources. This included functional annotations from the most recent 1000 Genomes Phase 3 baseline set of annotations (BaselineLD Version 2.2) recommended by the original developers of univariate S-LDSC.^17,54^ These annotations include minor allele frequency bins, histone marks, classes of enhancers and evolutionarily conserved variants, and flanking window annotations. We also include brain and endocrine relevant annotations for tissue specific gene expression from GTEx^55^and DEPICT^56^and histone marks from the Roadmap Epigenetics project.^57^ This was in addition to 5 randomly selected control regions for gene expression and histone marks (i.e., 10 controls total). Finally, we utilized 29 functional annotations that reflected the main effects of protein-truncating variant (PTV)-intolerant (PI) genes, human hippocampal and prefrontal brain cells, and their interactions. These annotations were created using collateral data from the Genome Aggregation Database (gnomAD)^58^ and GTEx^55^ data, with the parameters used to construct these annotations outlined in Grotzinger et al., 2020.

Flanking window, control, and continuous annotations were used to produce unbiased estimates of stratified genetic covariance but were excluded from enrichment analyses as these results would not be directly interpretable. This resulted in a total of 168 binary annotations. For SA, we removed 16 annotations that were non-positive definite and required smoothing the stratified covariance matrix such that any point estimate in the matrix produced a Z-statistic discrepancy > 1.96 pre- and post-smoothing. As we examined enrichment of the genetic variance for six factors (the five correlated factors and the general factor from the bifactor model) we employ a Bonferroni correction of *p* < 5.48E-5 (i.e., .05_FDR_/[152_annotations_ × 6_Genomic Factors_]). CT required removing 22 annotations due to high degrees of matrix smoothing, but we employ the same Bonferroni correction of *p* < 5.48E-5 for comparative purposes.

### Genetic Overlap with Cognitive Function

Genetic overlap with genetic *g* was examined in the context of the finalized bifactor and correlated factors models (i.e., those that included residual covariance across specific brain regions) for both CT and SA. This was achieved by simultaneously estimating all brain factor with *g*-factor correlations in the context of the models. As a first step in these analyses, we estimated an omnibus model chi-square difference test that compared the model fit for a model in which all correlations with the brain-based factors were freely estimated versus constrained to equality. This omnibus test asks at a general level whether *g* shows a uniform or factor-specific pattern of relationships for CT or SA. In line with the model fitting procedure used for the brain regions only models, residual covariances between individual brain regions and cognitive tests were iteratively added until they no longer reached a significance threshold of *p* < .01 within the context of the correlated factors model. For CT, this resulted in adding two residual covariances between verbal numerical reasoning and both the insula and precentral region. No residual covariances were added for SA. As each family of tests for CT and SA with *g* consisted of 204 possible associations (i.e., 34 brain regions × 7 cognitive tests) we employ a Bonferroni correction of .05/204 (*p* < 2.10E-4).

### Genetic Overlap with Psychiatric Disorders

The same four-factor, correlated factors model from Grotzinger et al. (2020)^19^ was used to model the multivariate architecture across the same 11 disorders. We note a few differences in the summary statistics used for the 11 disorders paper relative to the current analyses. This includes utilizing only publicly available summary statistics such that 23andMe data was not included for ADHD or MDD, using the most recent Freeze 3 summary statistics for bipolar disorder,^24^ and using the recently released anxiety summary statistics that reflect meta-analysis across the UK Biobank, ANGST, and iPSYCH consortium.^29^ We began by confirming that the model for psychiatric disorders only still provided good fit to the data even with the noted differences for some summary statistics (AIC = 162.86, CFI = .976, SRMR = .097), and that the factor loadings were largely concordant with those reported in prior work (Supplementary Table S10).

As with the *g-*factor analyses, we went on to examine the factor correlations across all psychiatric factors, and added residual covariance between individual disorders and brain regions until they no longer reached a significance threshold of *p* < .01. For SA, this resulted in adding in three residual relationships between autism spectrum disorder and the temporal pole, supra marginal, and rostral anterior cingulate regions. No residual covariances were identified for CT. We also estimated the same omnibus model chi-square difference test by comparing a model in which the correlations across the brain-based and psychiatric factors were fixed to equality versus freely estimates. As each family of tests for CT and SA with psychiatric disorders consisted of 374 possible associations (i.e., 34 brain regions × 11 disorders), we employed a Bonferroni correction of .05/374 (*p* < 1.34E-4).

## Data Availability

The data that support the findings of this study are all publicly available or can be requested for access. Specific download links for various datasets are directly below. 
Summary statistics for ENIGMA are available from: 
http://enigma.ini.usc.edu/research/download-enigma-gwas-results/
Summary statistics for the seven, individual cognitive traits are available from:
https://datashare.is.ed.ac.uk/handle/10283/3756
Summary statistics for data from the PGC can be downloaded or requested here: 
https://www.med.unc.edu/pgc/download-results/
Summary statistics for the Anxiety phenotype can be downloaded here: 
https://drive.google.com/drive/folders/1fguHvz7l2G45sbMI9h_veQun4aXNTy1v
Data from gnomAD used to identify PI genes for creation of annotations can be downloaded here: https://storage.googleapis.com/gnomad-public/release/2.1.1/constraint/gnomad.v2.1.1.lof_metrics.by_gene.txt.bgz
Gene count data per cell for creation of annotations were obtained from: https://storage.googleapis.com/gtex_additional_datasets/single_cell_data/GTEx_droncseq_hip_pcf.tar 
Data which maps individual cells to cell types (e.g. neuron, astrocyte etc.) were obtained from: https://static-content.springer.com/esm/art%3A10.1038%2Fnmeth.4407/MediaObjects/41592_2017_BFnmeth4407_MOESM10_ESM.xlsx
Links to the LD-scores, reference panel data, and the code used to produce the current results can all be found at: https://github.com/GenomicSEM/GenomicSEM/wiki
Links to the BaselineLD v2.2 annotations can be found here:
https://data.broadinstitute.org/alkesgroup/LDSCORE/

https://datashare.is.ed.ac.uk/handle/10283/3756

https://www.med.unc.edu/pgc/download-results/

https://drive.google.com/drive/folders/1fguHvz7l2G45sbMI9h_veQun4aXNTy1v

https://storage.googleapis.com/gnomad-public/release/2.1.1/constraint/gnomad.v2.1.1.lof_metrics.by_gene.txt.bgz

https://storage.googleapis.com/gtex_additional_datasets/single_cell_data/GTEx_droncseq_hip_pcf.tar

https://github.com/GenomicSEM/GenomicSEM/wiki

https://data.broadinstitute.org/alkesgroup/LDSCORE/

## Code Availability

GenomicSEM software (which now includes the *write*.*model* functionality for extensive sets of traits), is an R package that is available from GitHub at the following URL: https://github.com/GenomicSEM/GenomicSEM

Directions for installing and using the GenomicSEM R package can be found at: https://github.com/GenomicSEM/GenomicSEM/wiki

## Data Availability

The data that support the findings of this study are all publicly available or can be requested for access. Specific download links for various datasets are directly below.

Summary statistics for ENIGMA are available from: http://enigma.ini.usc.edu/research/download-enigma-gwas-results/

Summary statistics for the seven, individual cognitive traits are available from: https://datashare.is.ed.ac.uk/handle/10283/3756

Summary statistics for data from the PGC can be downloaded or requested here: https://www.med.unc.edu/pgc/download-results/

Summary statistics for the Anxiety phenotype can be downloaded here: https://drive.google.com/drive/folders/1fguHvz7l2G45sbMI9h_veQun4aXNTy1v

Data from gnomAD used to identify PI genes for creation of annotations can be downloaded here: https://storage.googleapis.com/gnomad-public/release/2.1.1/constraint/gnomad.v2.1.1.lof_metrics.by_gene.txt.bgz

Gene count data per cell for creation of annotations were obtained from: https://storage.googleapis.com/gtex_additional_datasets/single_cell_data/GTEx_droncseq_hip_pcf.tar

Data which maps individual cells to cell types (e.g. neuron, astrocyte etc.) were obtained from: https://static-content.springer.com/esm/art%3A10.1038%2Fnmeth.4407/MediaObjects/41592_2017_BFnmeth4407_MOESM10_ESM.xlsx

Links to the LD-scores, reference panel data, and the code used to produce the current results can all be found at: https://github.com/GenomicSEM/GenomicSEM/wiki

Links to the BaselineLD v2.2 annotations can be found here: https://data.broadinstitute.org/alkesgroup/LDSCORE/

## Acknowledgements

The work presented here was supported by a gift from the Tommy Fuss Fund. ADG was supported by NIH Grants R01MH120219 and RF1AG073593. We add that the current analyses would not have been possible without the enormous efforts put forth by the investigators and participants from ENIGMA, the Psychiatric Genetics Consortium, iPSYCH, and UK Biobank.

## Declaration of Interests

J.W.S. is a member of the Leon Levy Foundation Neuroscience Advisory Board, the Scientific Advisory Board of Sensorium Therapeutics, and has received honoraria for internal seminars at Biogen, Inc and Tempus Labs. He is PI of a collaborative study of the genetics of depression and bipolar disorder sponsored by 23andMe for which 23andMe provides analysis time as in-kind support but no payments.

---

**Table 1.** Summary of External Traits

| Cognitive Traits |  |  |  |  |
| --- | --- | --- | --- | --- |
| Trait | Data Source | Total Sample Size |  |  |
| Reaction Time<sup>18</sup> | UKB | 330,024 |  |  |
| Matrix pattern recognition<sup>18</sup> | UKB | 11,356 |  |  |
| Verbal numerical reasoning<sup>18</sup> | UKB | 171,304 |  |  |
| Symbol digit substitution<sup>18</sup> | UKB | 87,741 |  |  |
| Memory pairs-matching test<sup>18</sup> | UKB | 331,679 |  |  |
| Tower rearranging<sup>18</sup> | UKB | 11,263 |  |  |
| Trail making test-B<sup>18</sup> | UKB | 78,547 |  |  |
| Psychiatric Traits |  |  |  |  |
| Trait | Data Source | Cases | Controls | Population Prevalence |
| Alcohol use Disorder<sup>30</sup> | PGC | 8,485 | 20,272 | 15.90 |
| Anorexia Nervosa<sup>20</sup> | PGC | 16,992 | 55,525 | 0.90 |
| Anxiety Disorders<sup>29</sup> | UKB + ANGST + iPSYCH | 31,977 | 82,114 | 20.00 |
| Attention-deficit Hyperactivity Disorder<sup>26</sup> | PGC | 19,099 | 34,194 | 5.00 |
| Autism Spectrum Disorder<sup>25</sup> | PGC | 18,381 | 27,969 | 1.20 |
| Bipolar Disorder<sup>24</sup> | PGC | 41,917 | 371,549 | 2.00 |
| Major Depressive Disorder<sup>28</sup> | PGC + UKB | 170,756 | 329,443 | 15.00 |
| Obsessive Compulsive Disorder<sup>21</sup> | PGC | 2,688 | 7,037 | 2.50 |
| Post-traumatic Stress Disorder<sup>27</sup> | PGC | 2,424 | 7,113 | 30.00 |
| Schizophrenia<sup>23</sup> | PGC | 53,386 | 77,258 | 1.00 |
| Tourette's Syndrome<sup>22</sup> | PGC | 4,819 | 9,488 | 0.80 |

*Note.* The top half of the table reports information for the cognitive phenotypes while the bottom half reports information for the 11 psychiatric traits. Sample sizes reported are for the European ancestry only subsets of the summary statistics. We note that for many of the psychiatric summary statistics there were SNP-specific sum of effective sample sizes were provided, and that these effective sample sizes were used for relevant calculations within Genomic SEM. The population prevalence column lists the prevalence used for liability scale conversion. The broad depression phenotype was specifically used from the UKB Major Depression GWAS. The lifetime anxiety disorder phenotype was used for the UKB Anxiety GWAS. The PGC ALCH GWAS used the unrelated subsample from the overall genotyped European sample. We note that the sample sizes listed are the raw totals and do not reflect effective sample sizes.

## Online Supplement

**Supplementary Figure 1.**
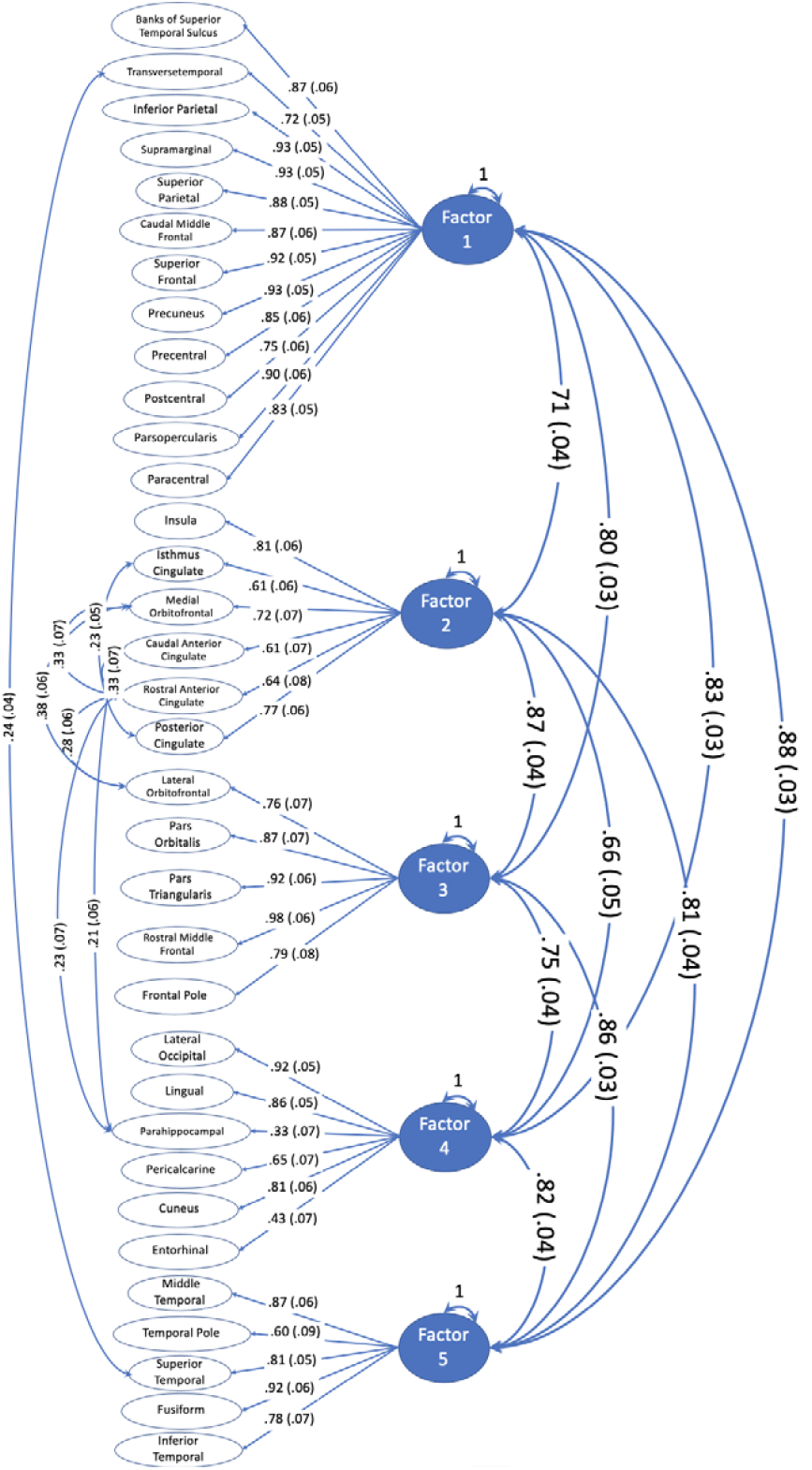
Correlated Factor Model Results for Cortical Thickness. Figure depicts the standardized loadings from the five-factor correlated factor model results for cortical thickness. All models used unit variance identification (i.e., factor variances were fixed to 1) and the associations between factors consequently depict genetic correlations. The residual associations across pairs of brain regions depict residual genetic correlations that are standardized relative to their SNP-based heritabilities (i.e., not standardized with respect to the residual genetic variance). The residual variances of the brain region indicators are not depicted for simplicity, but can be found in Supplementary Table 2.

**Supplementary Figure 2.**
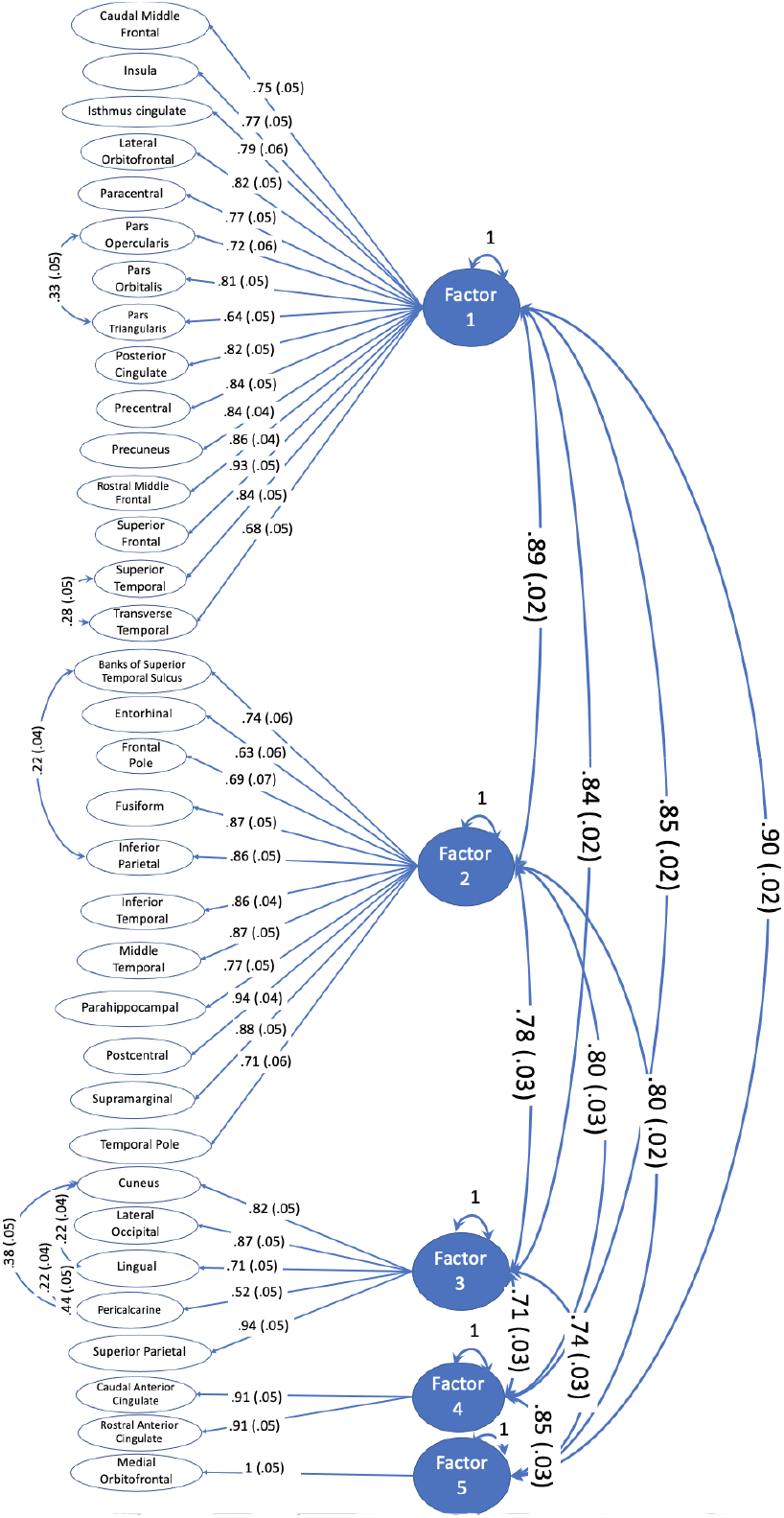
Correlated Factor Model Results for Surface Area. Figure depicts the standardized loadings from the five-factor correlated factor model results for Surface Area. All models used unit variance identification (i.e., factor variances were fixed to 1) and the associations between factors consequently depict genetic correlations. The bivariate associations between individual brain regions depict residual genetic correlations that are standardized relative to their SNP-based heritabilities (i.e., not standardized with respect to the residual genetic variance). The residual variances of the brain region indicators are not depicted for simplicity, but can be found in Supplementary Table 3.

**Supplementary Figure 3.**
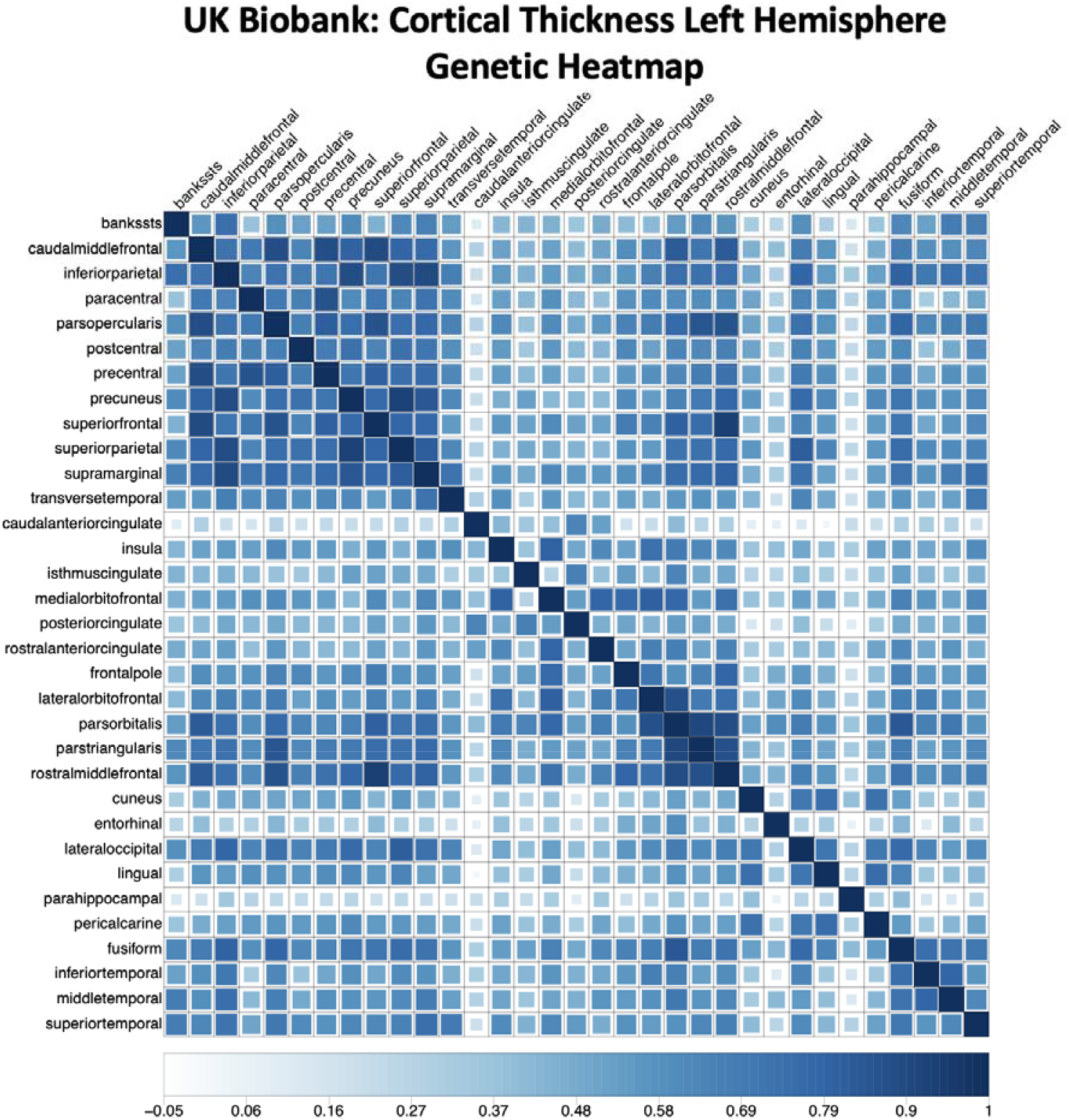
Genetic Heatmap for Left Hemisphere Cortical Thickness from UK Biobank. Figure depicts the genetic heatmap, as estimated using LD-score regression, for cortical thickness in the left hemisphere from UK Biobank. The heatmap is ordered with respect to the ENIGMA cortical thickness factor model identified in Genomic SEM.

**Supplementary Figure 4.**
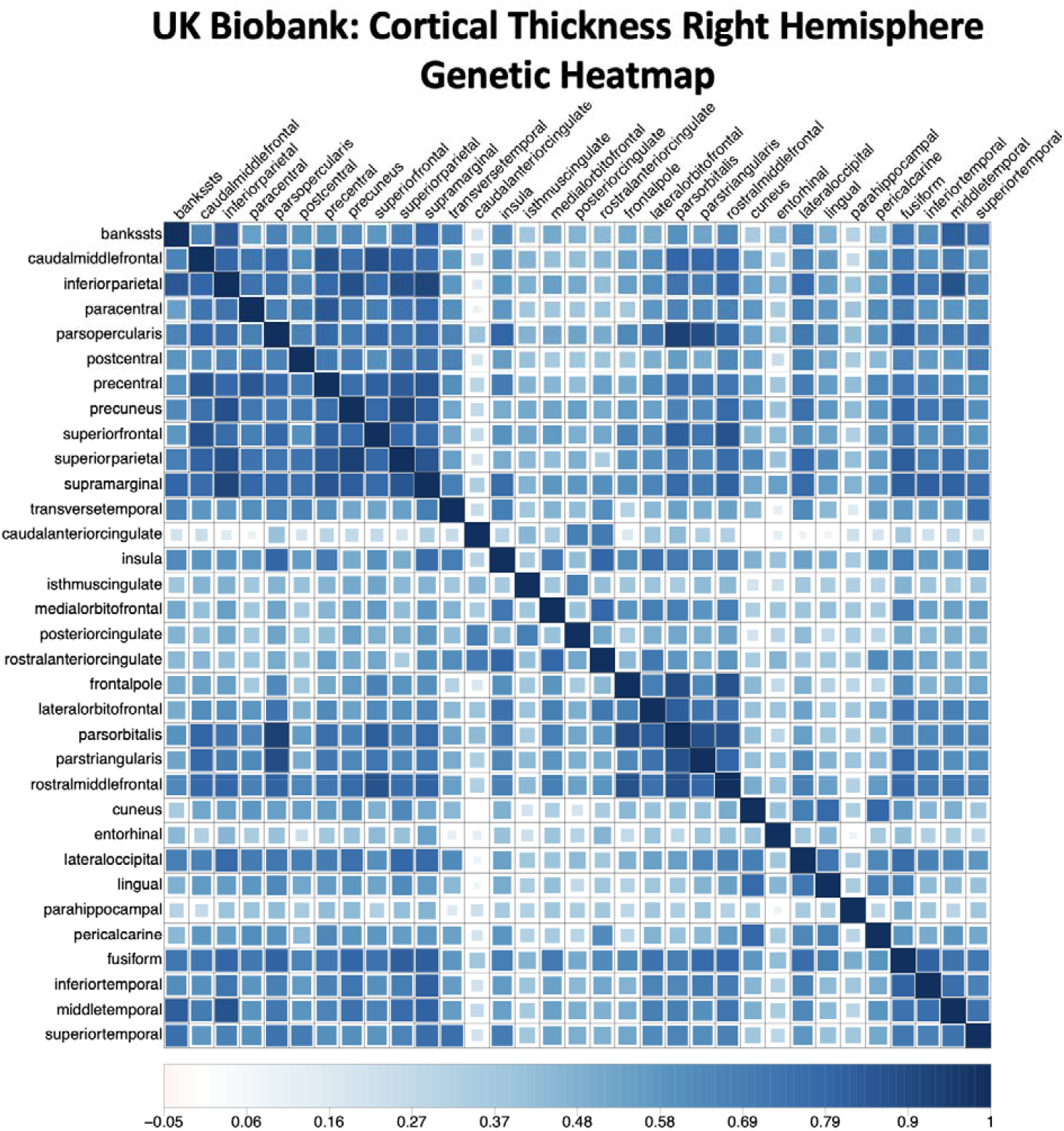
Genetic Heatmap for Right Hemisphere Cortical Thickness from UK Biobank. Figure depicts the genetic heatmap, as estimated using LD-score regression, for cortical thickness in the right hemisphere from UK Biobank. The heatmap is ordered with respect to the ENIGMA cortical thickness factor model identified in Genomic SEM.

**Supplementary Figure 5.**
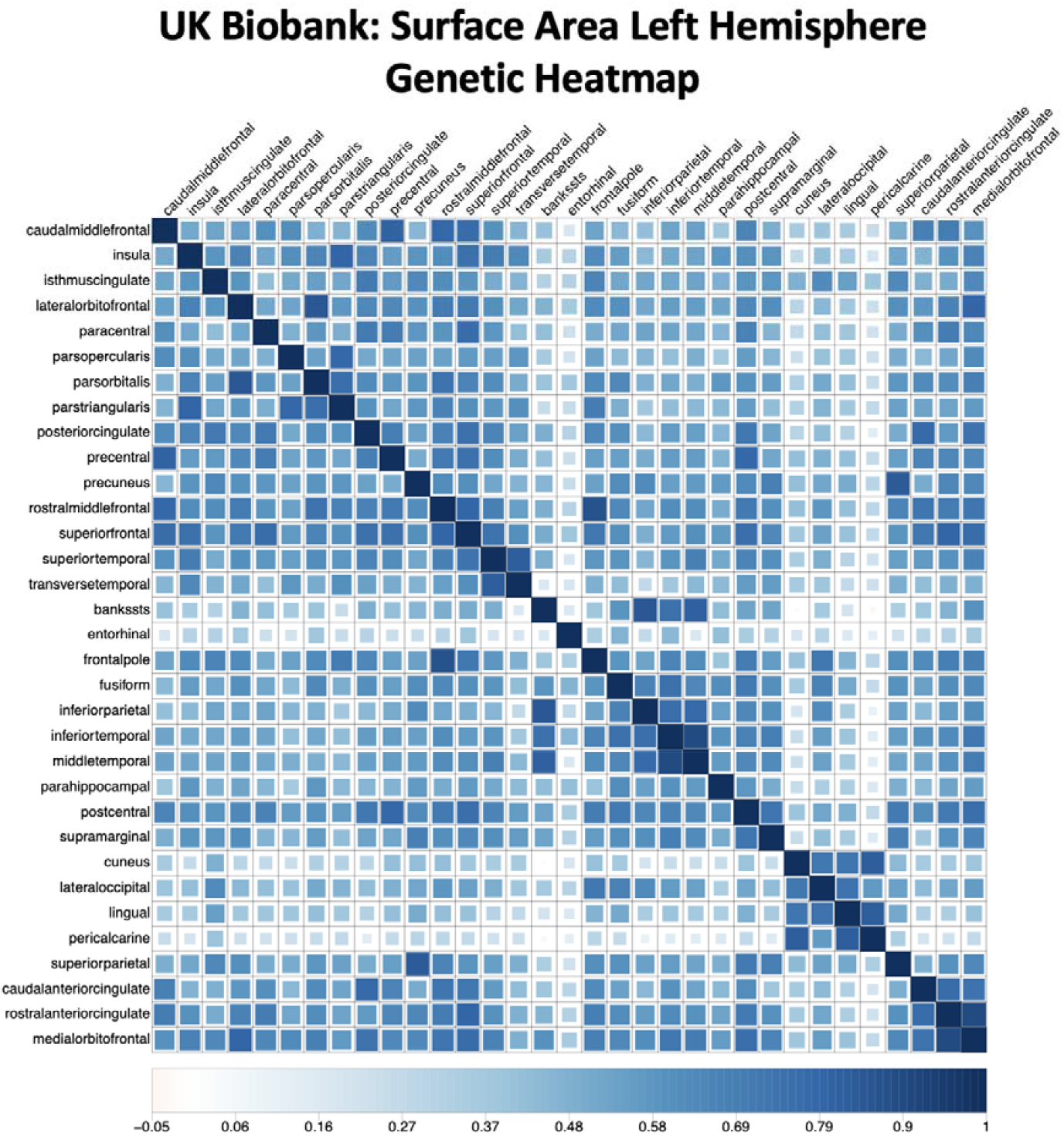
Genetic heatmap for Left Hemisphere Surface Area from UK Biobank. Figure depicts the genetic heatmap, as estimated using LD-score regression, for surface area in the left hemisphere from UK Biobank. The heatmap is ordered with respect to the ENIGMA surface area factor model identified in Genomic SEM.

**Supplementary Figure 6.**
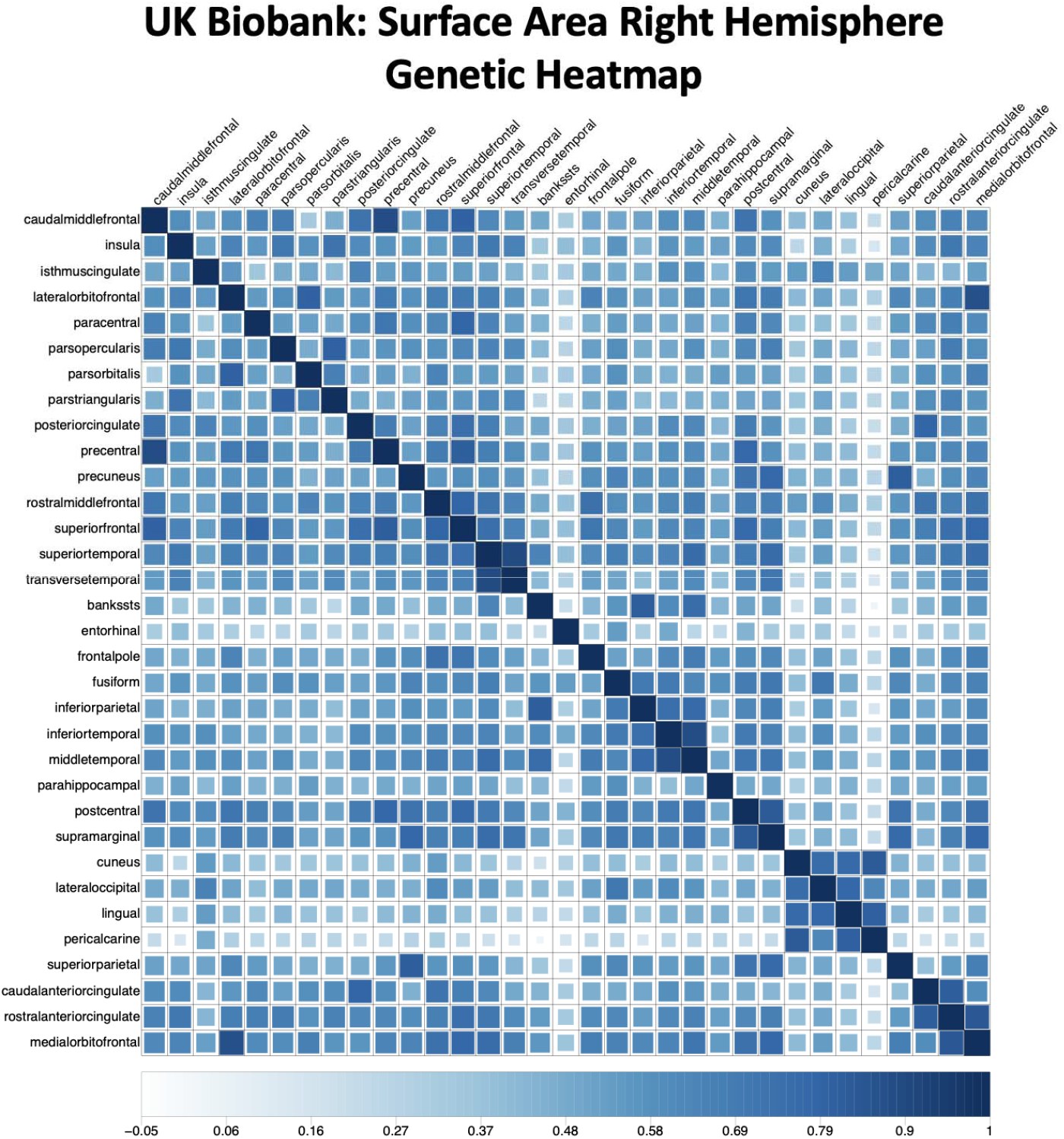
Genetic heatmap for Right Hemisphere Surface Area from UK Biobank. Figure depicts the genetic heatmap, as estimated using LD-score regression, for surface area in the right hemisphere from UK Biobank. The heatmap is ordered with respect to the ENIGMA surface area factor model identified in Genomic SEM.

**Supplementary Figure 7.**
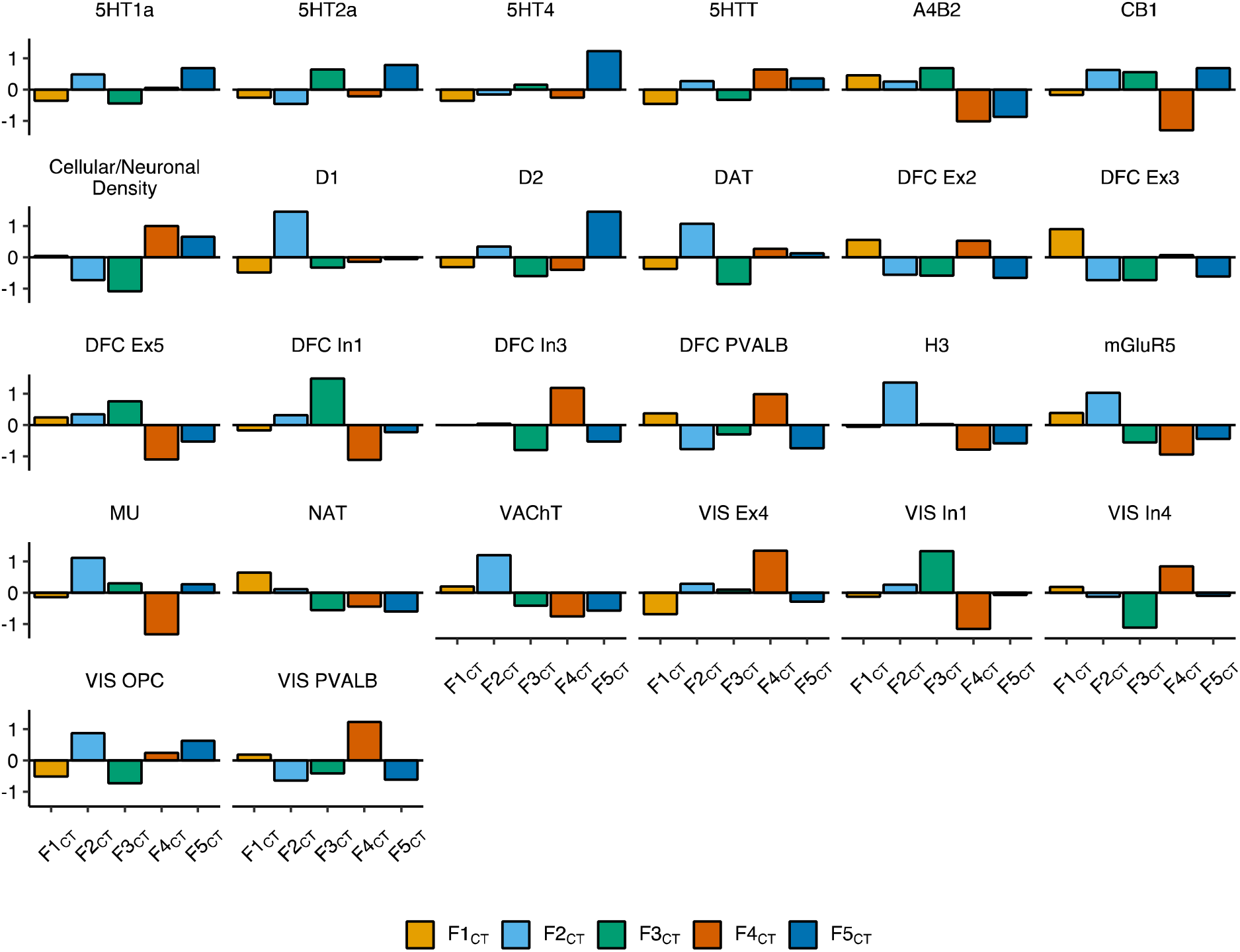
Biologically-derived features of the cortex that vary across the genomic factors of cortical thickness (CT). Faceted bar charts of mean standardized values per factor for biologically-derived features of the cortex. All displayed features were significantly different across the five genomic factors of CT (F1_CT_ to F5_CT_) in an omnibus test of differences (FDR-corrected *P*_spin_ < .05, Method). Full results are reported in Supplementary Table 9.

**Supplementary Figure 8.**
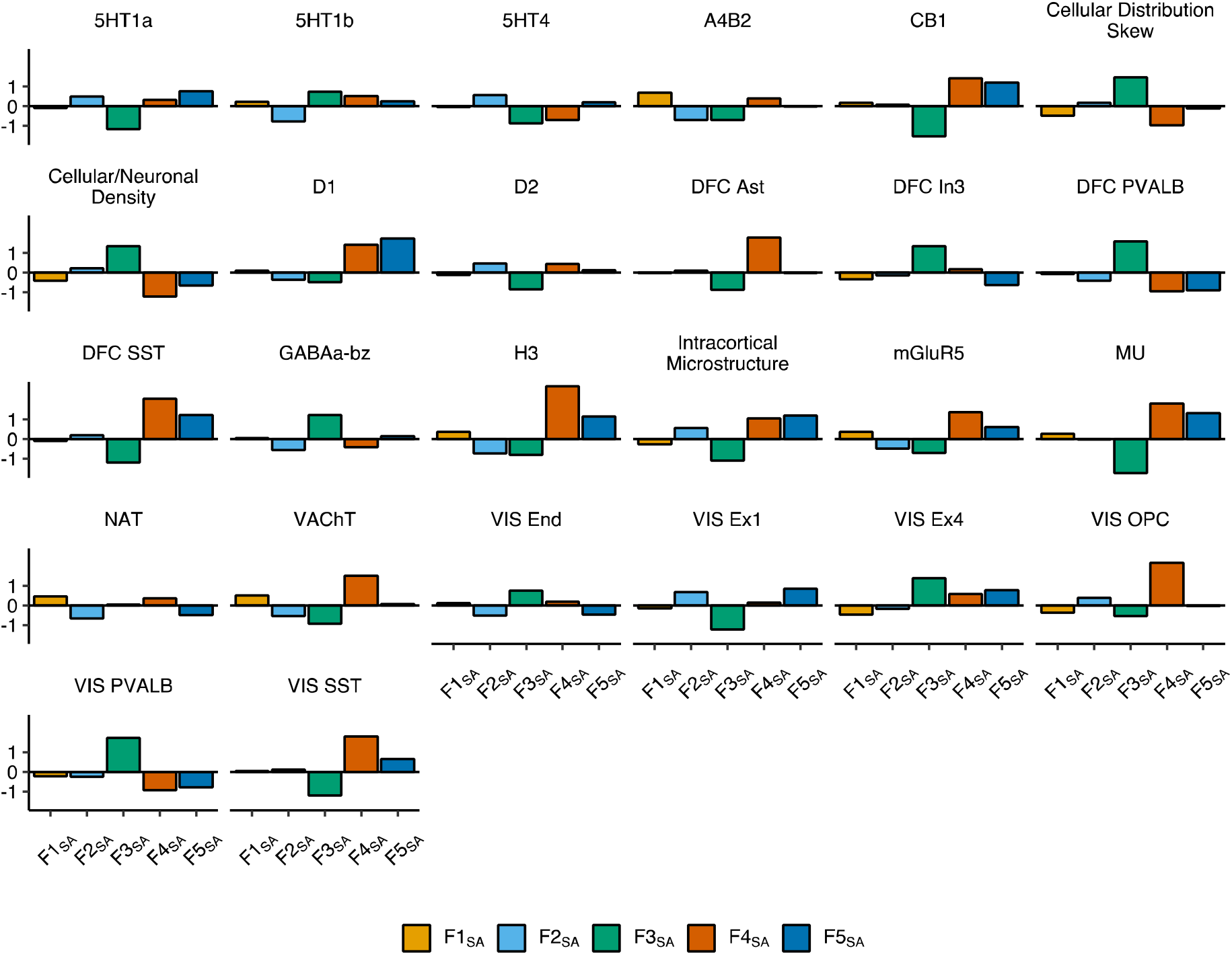
Biologically-derived features of the cortex that vary across the genomic factors of surface area (SA). Faceted bar charts of mean standardized values per factor for biologically-derived features of the cortex. All displayed features were significantly different across the five genomic factors of SA (F1_SA_ to F5_SA_) in an omnibus test of differences (FDR-corrected *P*_spin_ < .05, Method). Full results are reported in Supplementary Table 9.

**Supplementary Figure 9a.**
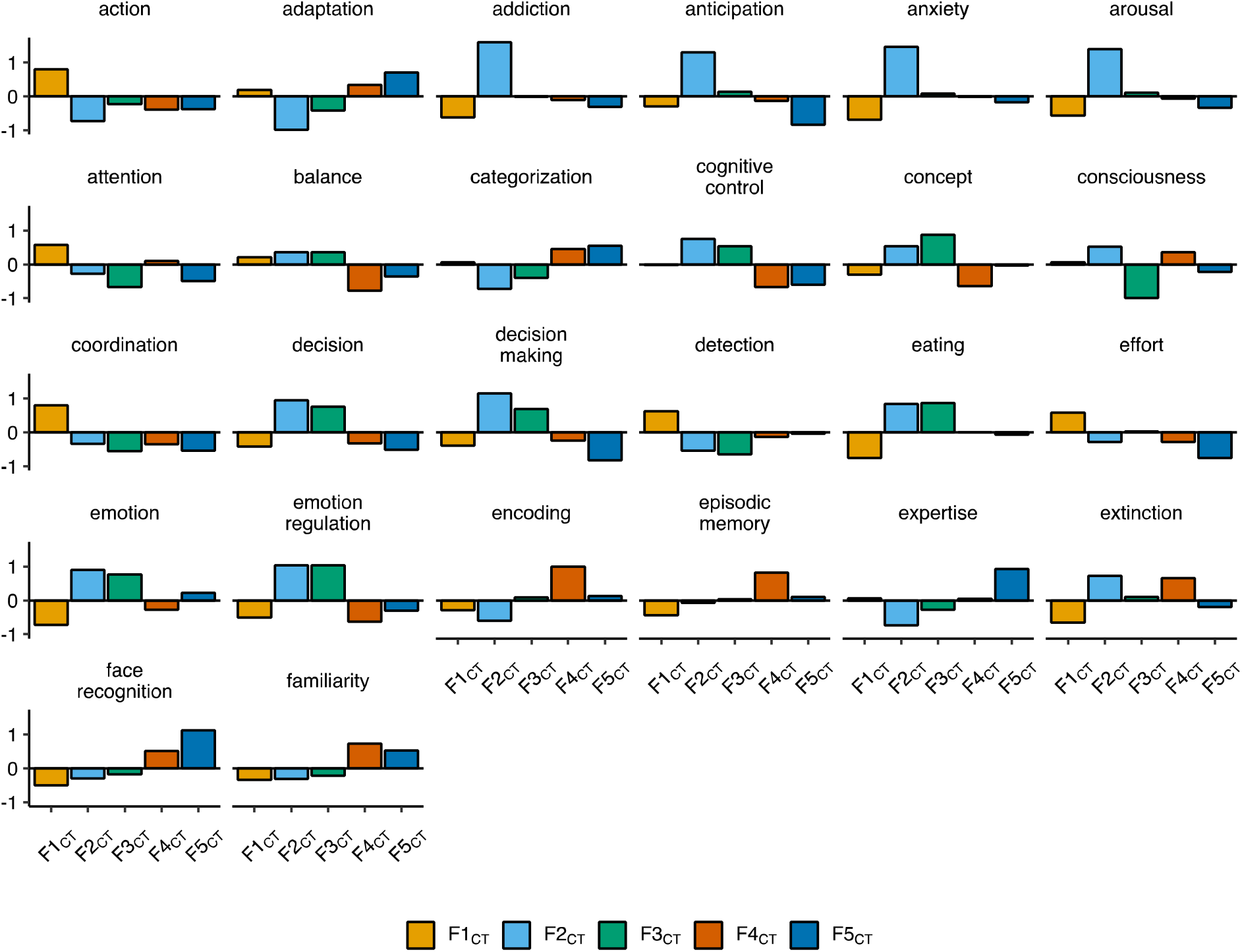
Functionally-derived features of the cortex that vary across the genomic factors of cortical thickness (CT). Faceted bar charts of mean standardized values per factor for functionally-derived features of the cortex. All displayed features were significantly different across the five genomic factors of CT (F1_CT_ to F5_CT_) in an omnibus test of differences (FDR-corrected *P*_spin_ < .05, Method). Full results are reported in Supplementary Table 9.

**Supplementary Figure 9b.**
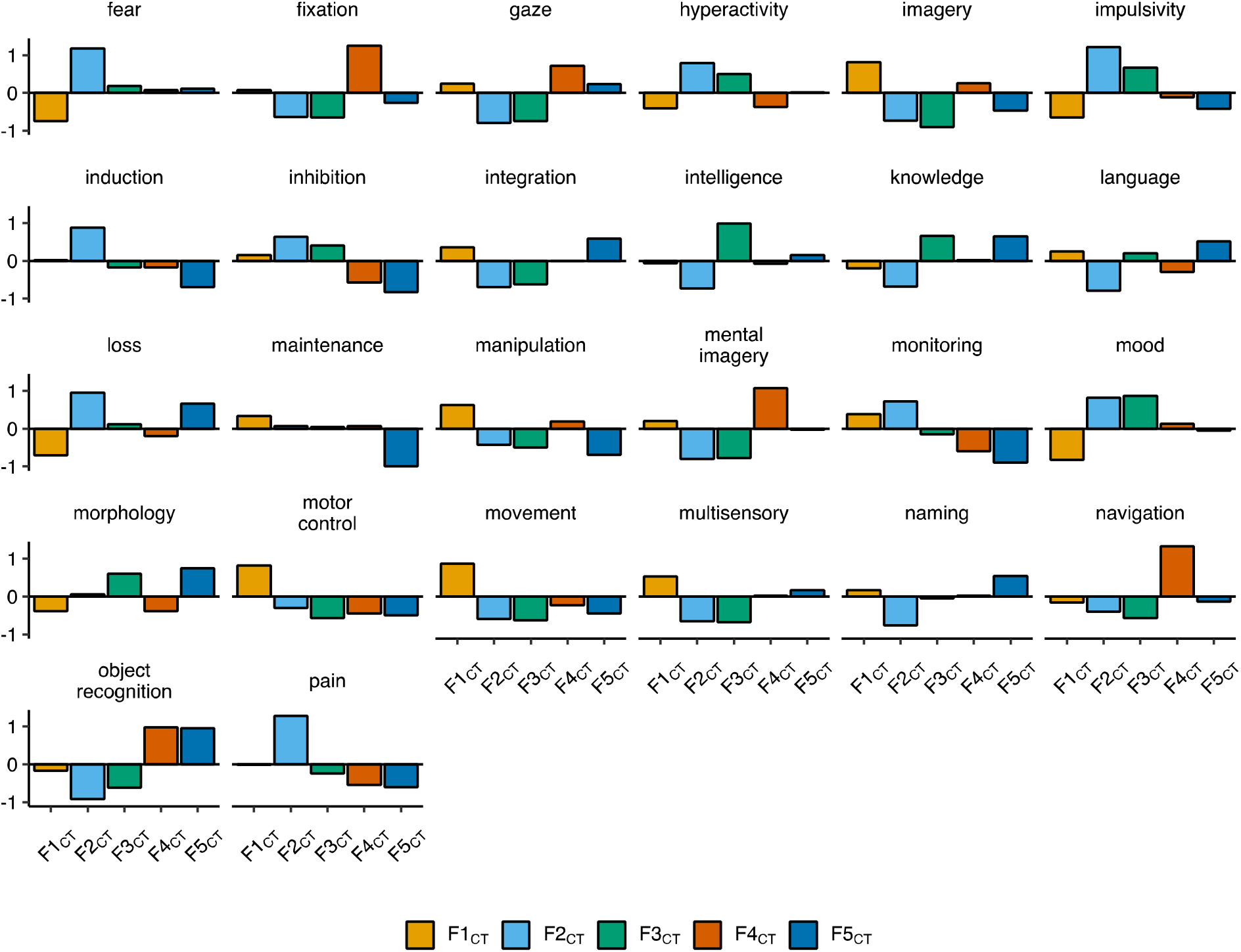
Functionally-derived features of the cortex that vary across the genomic factors of cortical thickness (CT). Faceted bar charts of mean standardized values per factor for functionally-derived features of the cortex. All displayed features were significantly different across the five genomic factors of CT (F1_CT_ to F5_CT_) in an omnibus test of differences (FDR-corrected *P*_spin_ < .05, Method). Full results are reported in Supplementary Table 9.

**Supplementary Figure 9c.**
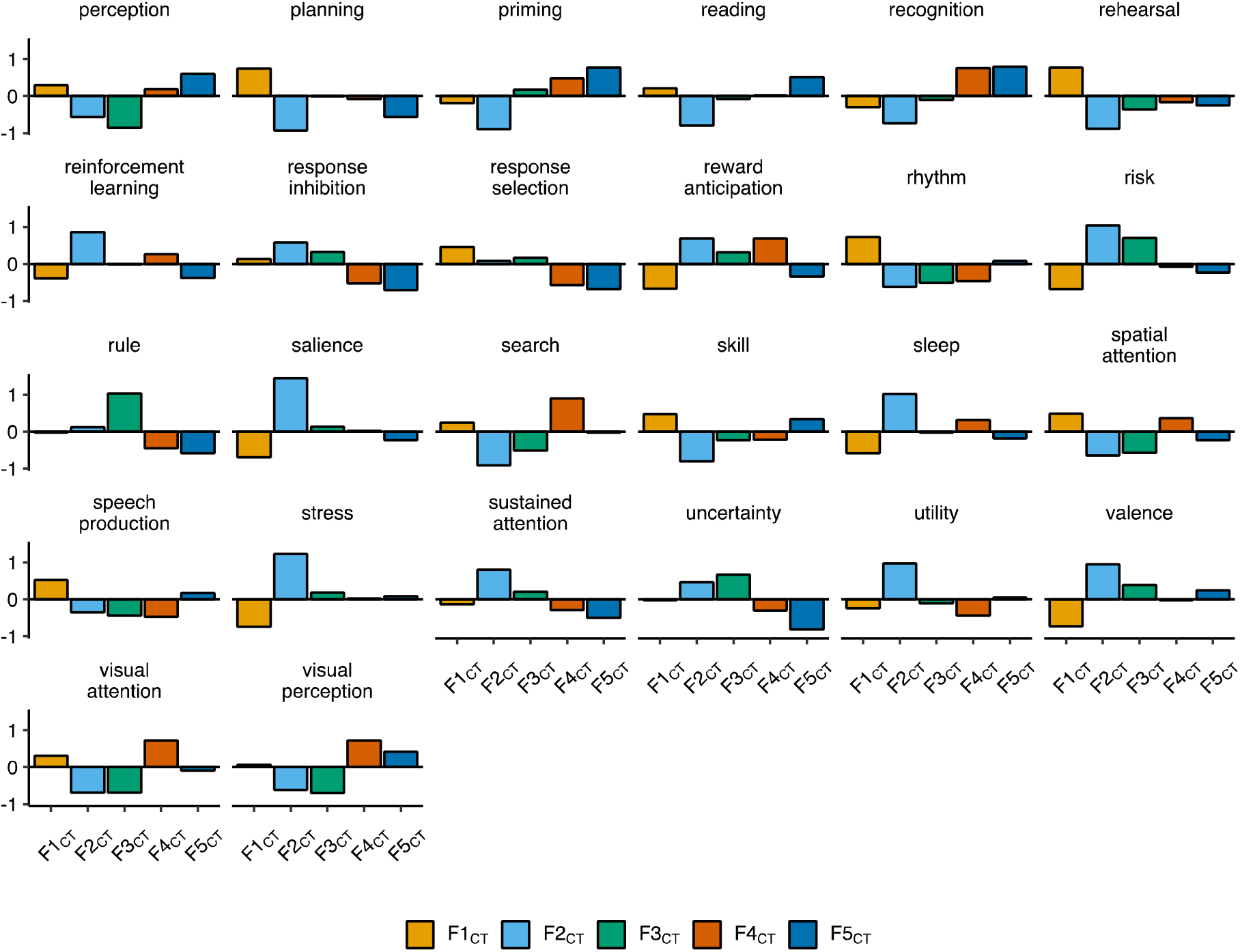
Functionally-derived features of the cortex that vary across the genomic factors of cortical thickness (CT). Faceted bar charts of mean standardized values per factor for functionally-derived features of the cortex. All displayed features were significantly different across the five genomic factors of CT (F1_CT_ to F5_CT_) in an omnibus test of differences (FDR-corrected *P*_spin_ < .05, Method). Full results are reported in Supplementary Table 9.

**Supplementary Figure 10a.**
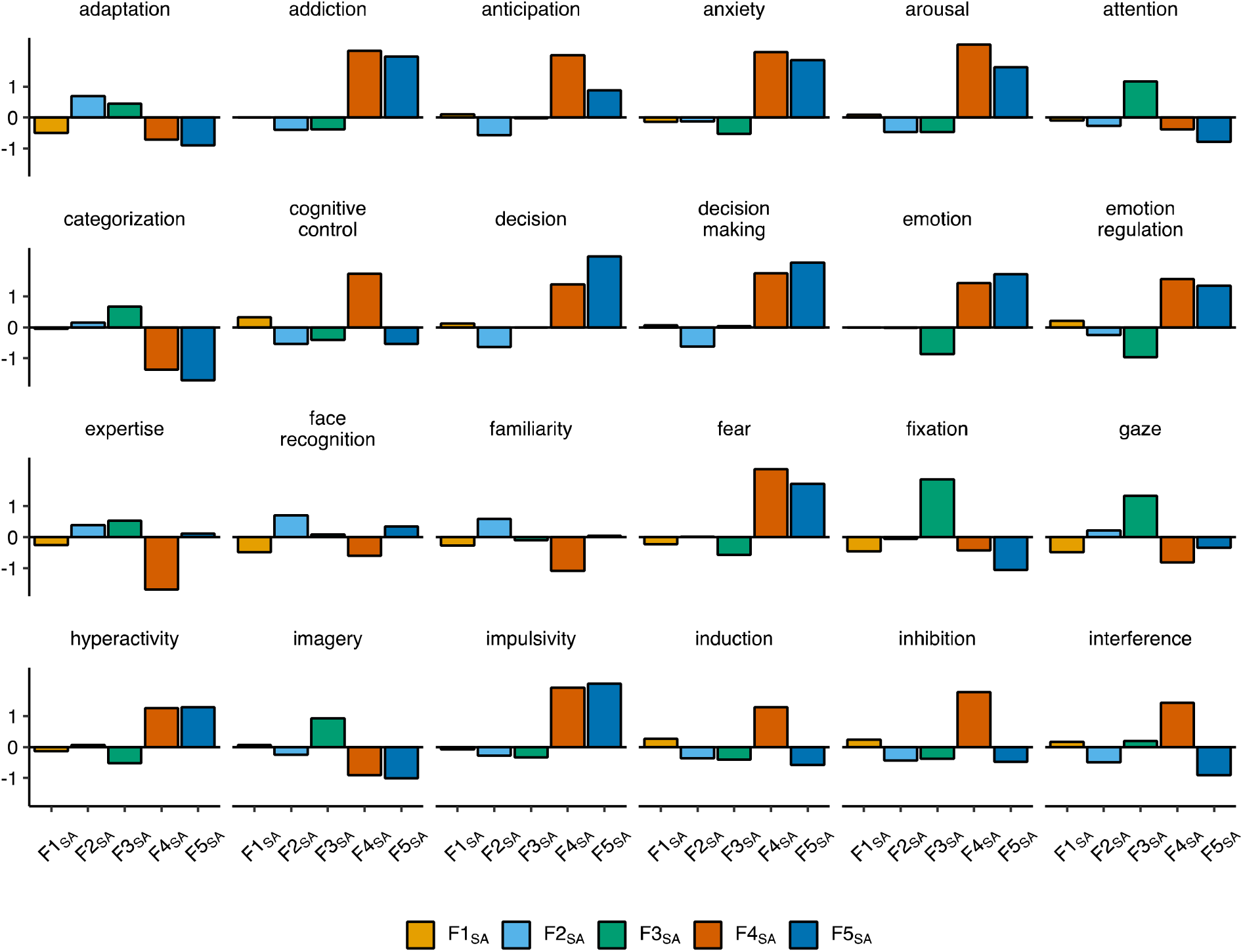
Functionally-derived features of the cortex that vary across the genomic factors of surface area (SA). Faceted bar charts of mean standardized values per factor for functionally-derived features of the cortex. All displayed features were significantly different across the five genomic factors of SA (F1_SA_ to F5_SA_) in an omnibus test of differences (FDR-corrected *P*_spin_ < .05, Method). Full results are reported in Supplementary Table 9.

**Supplementary Figure 10b.**
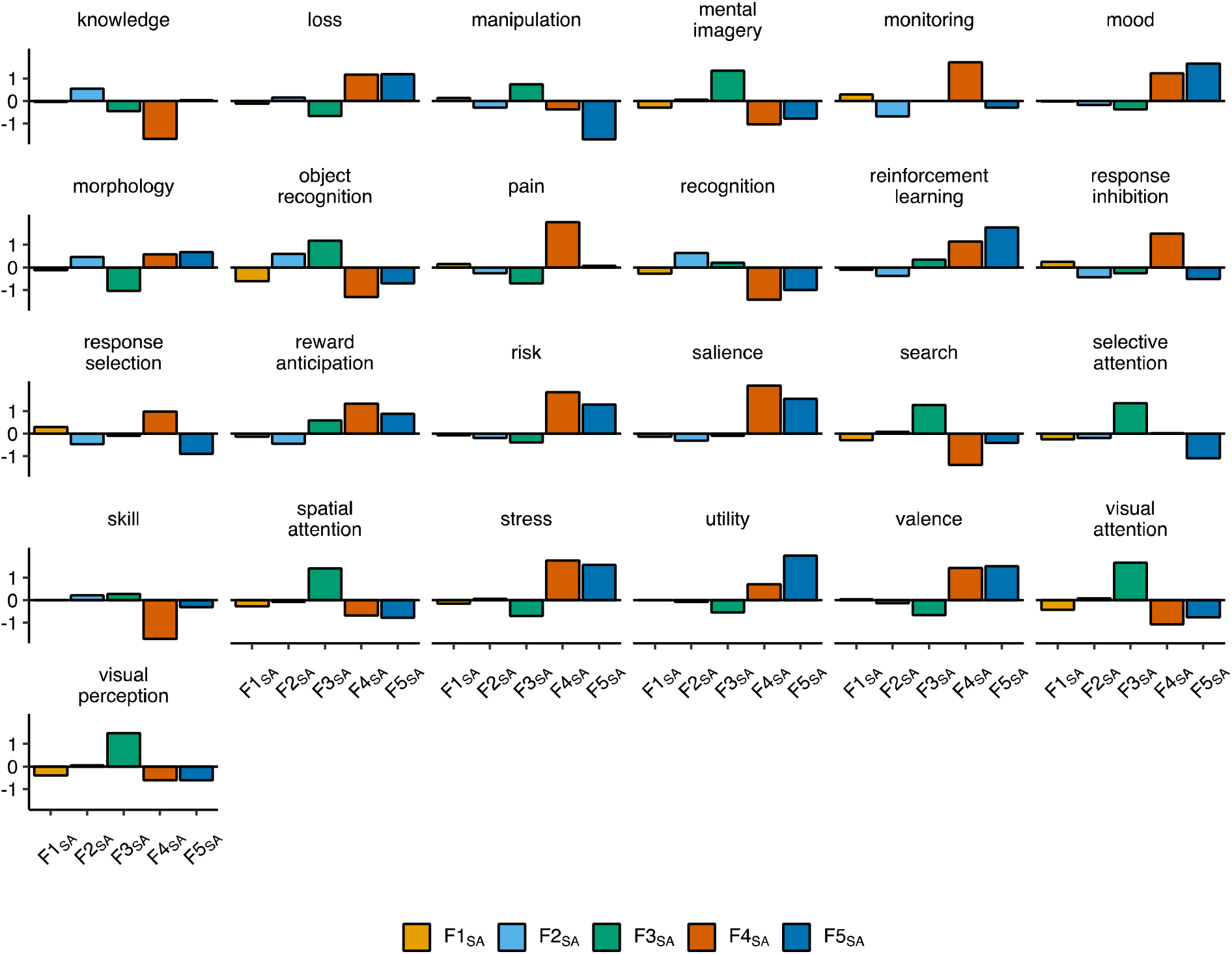
Functionally-derived features of the cortex that vary across the genomic factors of surface area (SA). Faceted bar charts of mean standardized values per factor for functionally-derived features of the cortex. All displayed features were significantly different across the five genomic factors of SA (F1_SA_ to F5_SA_) in an omnibus test of differences (FDR-corrected *P*_spin_ < .05, Method). Full results are reported in Supplementary Table 9.

## Footnotes

1 *Given the large number of variables used in this analysis, coupled with the expanding landscape of GWAS summary statistics available, we have tested, coded, and made publicly available a new function within the GenomicSEM R package*, ***write*.*model***, *that will automatically generate the CFA model syntax based on output from an EFA*.

## Notes

### Author Declarations

The data that support the findings of this study are all publicly available or can be requested for access. Specific download links for various datasets are directly below. Summary statistics for ENIGMA are available from: http://enigma.ini.usc.edu/research/download-enigma-gwas-results/ Summary statistics for the seven, individual cognitive traits are available from: https://datashare.is.ed.ac.uk/handle/10283/3756 Summary statistics for data from the PGC can be downloaded or requested here: https://www.med.unc.edu/pgc/download-results/ Summary statistics for the Anxiety phenotype can be downloaded here: https://drive.google.com/drive/folders/1fguHvz7l2G45sbMI9h_veQun4aXNTy1v Data from gnomAD used to identify PI genes for creation of annotations can be downloaded here: https://storage.googleapis.com/gnomad-public/release/2.1.1/constraint/gnomad.v2.1.1.lof_metrics.by_gene.txt.bgz Gene count data per cell for creation of annotations were obtained from: https://storage.googleapis.com/gtex_additional_datasets/single_cell_data/GTEx_droncseq_hip_pcf.tar Data which maps individual cells to cell types (e.g. neuron, astrocyte etc.) were obtained from: https://static-content.springer.com/esm/art%3A10.1038%2Fnmeth.4407/MediaObjects/41592_2017_BFnmeth4407_MOESM10_ESM.xlsx Links to the LD-scores, reference panel data, and the code used to produce the current results can all be found at: https://github.com/GenomicSEM/GenomicSEM/wiki Links to the BaselineLD v2.2 annotations can be found here: https://data.broadinstitute.org/alkesgroup/LDSCORE/

### Summary of Updates

The formatting was shifted down such that table and figure notes were spread across multiple pages. This revision is being submitted strictly to update these formatting issues.

